# Possible roles of phytochemicals with bioactive properties in the prevention of and recovery from COVID-19

**DOI:** 10.1101/2024.01.29.24301882

**Authors:** Sachiko Koyama, Vonnie D.C. Shields, Thomas Heinbockel, Paule V. Joseph, Poonam Adhikari, Rishemjit Kaur, Ritesh Kumar, Rafieh Alizadeh, Surabhi Bhutani, Orietta Calcinoni, Carla Mucignat-Caretta, Jingguo Chen, Keiland W. Cooper, Subha R. Das, Paloma Rohlfs Domínguez, Maria Dolors Guàrdia, Maria A. Klyuchnikova, Tatiana K. Laktionova, Eri Mori, Zeinab Namjoo, Ha Nguyen, Mehmet Hakan Özdener, Shima Parsa, Elif Özdener Poyraz, Daniel Jan Strub, Farzad Taghizadeh-Hesary, Rumi Ueha, Vera V. Voznessenskaya

## Abstract

**Purpose:** There have been large geographical differences in the infection and death rates of COVID-19. Foods and beverages containing high amounts of phytochemicals with bioactive properties were suggested to prevent contracting, to limit the severity of, and to facilitate recovery from COVID-19. The goal of our study was to determine the correlation of the type of foods/beverages people consumed and the risk reduction of contracting COVID-19 and the recovery from COVID-19.

**Methods:** We developed an online survey that asked the participants whether they contracted COVID-19, their symptoms, time to recover, and their frequency of eating various types of foods/beverages. The survey was first developed in English and then translated into 10 different languages.

**Results:** The participants who did not contract COVID-19 consumed vegetables, herbs/spices, and fermented foods/beverages significantly more than the participants who contracted COVID-19 and those who were not tested but became sick most likely from COVID-19. The geographic location of participants corresponded with the language of the survey, except for the English version, thus, nine out of the 10 language versions represented a country. Among the six countries (India, Iran, Italy, Japan, Russia, Spain) with over one hundred participants, we found that in India and Japan the people who contracted COVID-19 showed significantly shorter recovery time, and greater daily intake of vegetables, herbs/spices, and fermented foods/beverages was associated with faster recovery.

**Conclusion:** Our results suggest that phytochemical compounds included in the vegetables may have contributed in not only preventing contraction of COVID-19, but also accelerating their recovery. (249 words; EJN limit is 250 words)

## Introduction

The outbreak of the coronavirus disease 2019 (COVID-19) became a long pandemic that lasted over three years, and, although the federal COVID-19 public health emergency declaration ended in 5/11/2023, there are still new variants of the SARS-CoV-2 prevailing and weekly over 30,000 new patients in the U.S were hospitalized in January 2024 (https://covid.cdc.gov/covid-data-tracker/#datatracker-home). As of January 7th, 2024, the cumulative cases of patients who contracted the virus surpassed 774 million with over 7 million deaths worldwide (https://data.who.int/dashboards/covid19/cases?n=c). Throughout the pandemic, there have been various reports investigating the factors that may influence infection, the severity of symptoms, and the recovery process including genetic factors, pre-existing conditions, and age. We now know that the polymorphism of ACE2, the genes that regulate ACE2 expression and affinity to SARS-CoV-2, and that sex affect the susceptibility to COVID-19 (for example, [1–5]). Pre-existing conditions such as obesity and type 2 diabetes [5, 6] as well as age [5] have also been found to affect vulnerability. Another key point that also emerged from previous studies was the geographical difference in the number of cases around the world. Many speculated that it may have been the result of government strategies, such as herd immunity, and/or genetic differences [7, 8]. There were also discussions around the possibility of differences in the food materials used by different countries and the effects of phytochemical compounds included in them contributing to prevent contracting the virus and/or improving the recovery process [9].

Since the outbreak of COVID-19, there have been multiple studies examining the effects of phytochemicals and plant-based diets and beverages on COVID-19 (e.g., [10, 11] on tea; [12, 13] on plant-based diet; [14] on the effects of eating carnation and citrus fruits). *In silico* and review studies have shown that there are phytochemicals which may bind to the SARS-CoV-2 responsible for COVID-19 (e.g., [15, 16]). Such binding may contribute to suppress the infection if the phytochemicals bind to the spike protein of the virus [9, 17], and to suppress the replication if they bind to the enzymes involved in the replication of the virus [9]. Foods that contain large amounts of phytochemical compounds with bioactive properties may resemble the effects of drugs used for treatment of COVID-19 [18]. These studies suggest that daily consumption of foods and beverages that contain these phytochemicals may prevent/suppress the contraction of COVID-19 or facilitate faster recovery from it [18–22]. Some examples of these foods and beverages are green vegetables, such as broccoli/cabbage, citrus fruits, such as lemon and orange and red fruits, such as blueberries, coffee, and black tea. Considering the studies indicating that some of the patients with severe symptoms for long term after the acute phase of COVID-19 (post-acute sequelae of COVID-19 (PASC), or Long-COVID) possess virus persisting in their tissue as a reservoir [23], consuming foods/beverages with such effects in their daily may have a possibility to be some help in their recovery by binding to the S-protein or the enzymes related to replication of virus.

Other than the effects of phytochemicals on binding to SARS-CoV-2 and limiting infection, there are phytochemicals with anti-inflammatory effects [15, 24]. As inflammation is one of the key symptoms of COVID-19 and PASC, phytochemicals with anti-inflammatory effects may have beneficial influences on the recovery from COVID-19 and PASC [25, 26].

There are countries where cuisines and beverages widely use various herbs and spices. Many of the phytochemicals included in these cuisines possess anti-inflammatory and antioxidant effects (for example zingerone of ginger [27, 28] and curcumin of turmeric [28, 29]), and some may bind to the SARS-CoV-2 that causes COVID-19 as well (for example carvacrol in oregano and thyme [17, 30], cinnamaldehyde in cinnamon [30], geraniol in lemongrass, lavender, rose and others [30], curcumin in turmeric [30–32] and others [33]). It is possible that depending on the traditional cuisines in each country, there are differences in the type and amount of these phytochemicals consumed, and that may have produced the differences in the infection, severity, and recovery from COVID-19. These phytochemicals are also included in beverages, such as teas from tea plants [10, 21] and herbal teas. A recent study conducted with healthy identical twins has shown that those who were assigned to consume vegan diet had significantly improved cholesterol concentration, fasting insulin level, and body weight loss compared to those who were assigned to eat omnivorous food, indicating the effects of phytochemicals on several health conditions even among genetically identical twins [34]. The health benefits of fermented foods has also been shown [35–37], which not only affect gut microbiota by the probiotic lactic acid bacteria, but also can improve the health status of the person directly by their anti-inflammatory, antioxidant, and other bioactive properties. Herbal medicines [38], supplements [39], and essential oils [40] may also function to prevent and improve the recovery from COVID-19 due to the phytochemicals with bioactive properties included in them.

The goal of the present study was to determine if there were associations between the contraction of COVID-19 and/or recovery from it and the self-reported consumption of phytochemicals. To do so, we developed a global survey to understand the possible correlation between the types and frequencies of the foods, beverages, herbal medicine, supplements, and essential oils people consumes or used, and COVID-19 infection rates and recovery. The results of our survey suggested that the participants of the survey who did not get COVID-19 ate leaf vegetables, root vegetables, other vegetables, and fermented foods/beverages, and used herbs and spices daily at a high percentage. In addition, the participants who contracted COVID-19 who answered that they ate vegetables, fermented foods/beverages, and used herbs and spices daily showed shorter days to recover from COVID-19.

## Materials and Methods

### Participants

Data was collected from participants who consented to participate and were older than 18 years in age. Participation in this survey was voluntary and was not remunerated. As it was an online survey, the participants had to access it via the internet. The protocol was approved as an exempt study by Indiana University Human Research Protection Program (HRPP) in the U.S.A. (protocol #14915) as well as by the Office of Regulatory Research Compliance of Howard University (IRB-2022-0380), by the Jikei University School of Medicine Ethics Committee in Japan (#34-003), the Bioethics Committee at the A.N. Severtsov Institute of Ecology & Evolution of Russian Academy of Sciences (no.2022-63-NC), and by the Second Affiliated Hospital of Xi’an Jiaotong University, Medical Ethics Committee in China (#2022023). The link and the QR codes to the survey were distributed using social media (for example, Twitter, Facebook, etc.), virtual communication platforms, flyers, and word of mouth. This study was conducted as an Internal Study of the Global Consortium for Chemosensory Research (GCCR) (https://gcchemosensr.org/), Project ID #NDS005, and invitation to participate in the survey was also sent to participants of previous survey by GCCR, Project ID #GCCR001 [41].

### Survey development

In the survey, we gathered information on demographic data, dietary habits, and information on COVID-19 infection experiences. The questions asked included: 1) age, 2) ethnicity, 3) location of residency, 4) experience of COVID-19 contraction, 5) symptoms experienced and severity, 6) eating and drinking habit and frequency of intake (daily, weekly, monthly, and never), 7) use of herbal medicine and frequency, 8) use of supplements and frequency, and 9) use of essential oils for prevention and recovery from COVID-19. The questions on foods and beverages were conducted in categorical questions and free text questions so that participants could write the name of the foods and beverages that were not in the list of foods and beverage examples shown in the survey. Considering the goals of the survey, we also asked whether a specific culture or a country influenced the answers to the questions on eating and drinking habits. Smoking habits and pre-existing conditions were also asked. Details on the survey development can be found in Supplementary Information.

### Processing and analyses of the results

All the data from each language version were combined in one file in order to obtain the “Overall results” (see the Results section). The results displayed the amount of time that it took individuals to complete the survey. Data were excluded if the time to complete the survey was short from the concern that the participant may have not taken the time needed to answer each survey question thoughtfully and accurately. We arbitrarily selected 100 seconds as the minimum time to finish the surveys. The participants who claimed to be younger than 18 years old were excluded. The participants who did not answer the question about whether they got COVID-19 were also excluded. Following this step, the distribution of selections of the frequency of eating and drinking (daily, weekly, monthly, never) and free text answers, depending on health condition (“Not tested”, “Tested positive”, and “Did not get COVID”), were analyzed.

Secondly, the data of each language version were analyzed separately. The answers concerning ethnicity and current location were analyzed to determine if each language version reflected any specific country and whether there was a specific country or culture affected. Subsequently, the distribution of selections of the frequency of eating and drinking (daily, weekly, monthly, never) and free text answers, depending on the health condition (“Not tested”, “Tested positive”, and “Did not get COVID”), were analyzed.

### Statistical analyses

We used Chi-square tests for comparisons of the food and beverage consumption, as well as the severity of symptoms. We used the Kruskal Wallis and Mann-Whitney U-tests to compare the days to recover from COVID-19. Systat 13.2 (Inpixon HQ, Palo Alto CA, USA) was used for the statistical analysis.

## Results

### Overall results (all countries together)

#### i) Number of participants used for data analyses

The survey was conducted online. Overall, there were 1777 participants. The time taken by participants to complete an online survey was interpreted to be a representation of their level of comprehension and accuracy in responding to the questions. To ensure the reliability of the collected data, we conducted an analysis of the survey completion times and found that about 10% of the participants (n=187) completed the survey within a significantly short period, *i.e*., within 100 sec, which suggest a lack of comprehension. We decided to exclude the participants who finished the survey within 100 seconds. Participants who were younger than 18 as well as the participants who did not answer the question on whether they contracted COVID-19 were also excluded. This made the amount of data from 1583 participants eligible to be used for further analysis.

#### ii) Gender and age of participants

The gender and age of the participants whose data were used in the analyses were male (average age 40.27±14.60; n=237 (26.87%)), female (average age 42.43±14.07; n=624 (70.75%)), non-binary/third gender (average age 37.50±13.96; n=4 (0.454%)), and prefer not to answer (average age 41.82±12.70, n=17 (1.51%)).

#### iii) COVID-19 status of the participants

The participants were asked about their COVID-19 status and to select from three categories, *i.e*. “Not tested but suspected”, “Tested positive”, and “Did not get COVID”. Of the 1583 participants, 312 participants (19.7 %) reported to not have been tested but suspected that they contracted COVID-19, 835 participants (52.7 %) reported that they had tested positive for the virus, and 436 participants (27.5%) reported to have not contracted COVID-19 or contracted it unknowingly.

#### iv) Symptoms

Overall, the participants of the “Tested positive” group reported more severe symptoms than the “Not tested” group. The symptoms of “Headache”, “Lightheadedness”, “Diarrhea”, “Memory loss” were significantly more reported by the “Tested positive” group (Chi-square test, Headache, χ2=20.244, df=4, P<0.001; Lightheadedness, χ2=19.026, df=4, P<0.001; Memory loss, χ2=20.993, df=4, P<0.001; Diarrhea, χ2=12.372, df=4, P<0.05) (Figure 1, Supplementary Table 2). There were no significant differences in other symptoms asked in the survey, such as fever, chills, body aches, and so on between the “Not tested” and “Tested positive” groups (Supplementary Figure 1, Supplementary Table 2).

**Fig. 1.**
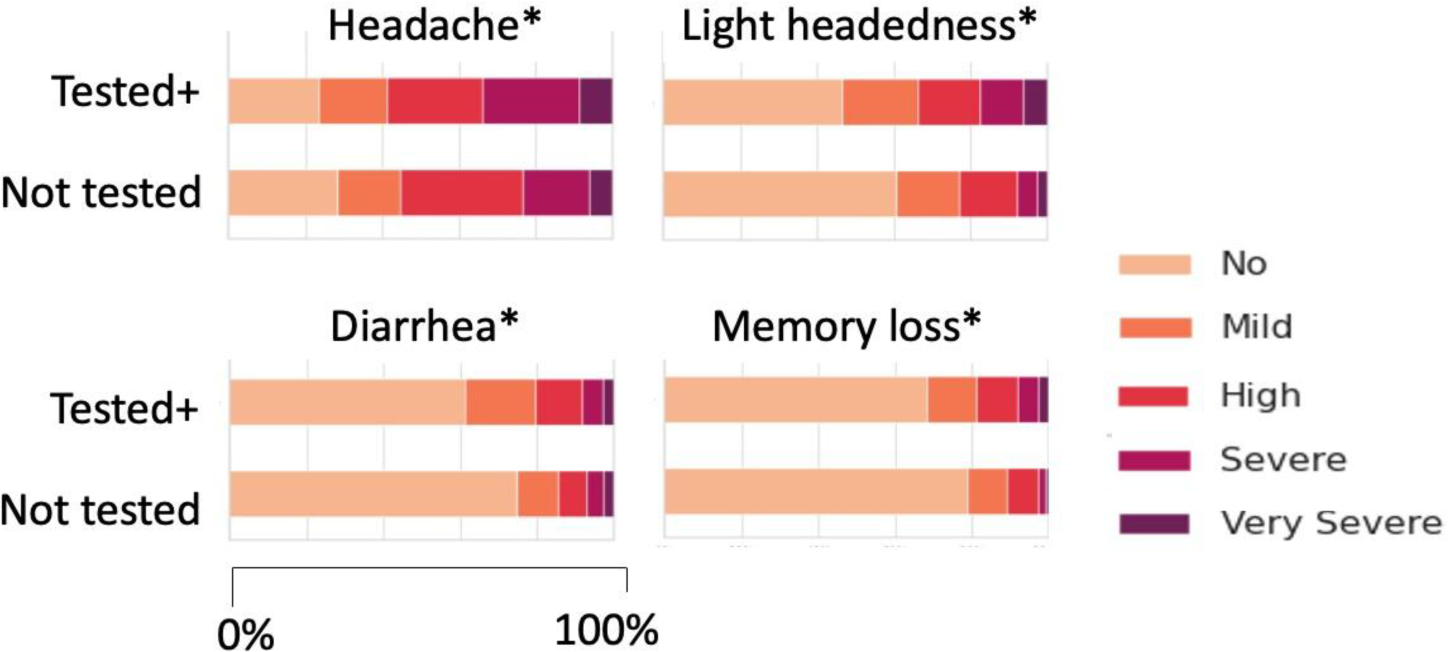
Symptoms that showed significant differences between the “Not tested” and “Tested positive” groups. *: Chi-square test, P<0.05.

Considering that the impact on chemical senses is known to be one of the major symptoms of COVID-19, we compared the self-reported dysfunction in the chemical senses. The senses of smell and taste were reported more frequently compared to chemesthesis, and there were no significant differences between the “Not tested” and “Tested positive” groups (Chi-square test, smell, χ2=2.122, df=2, N.S.; taste, χ2=4.013, df=2, N.S. Figure 2). Dysfunction in chemesthesis was less reported compared to the senses of smell and taste, and there were no differences between the “Not tested” and “Tested positive” group (Chi-square test, χ2=2.532, df=2, N.S.; Figure 2, Supplementary Table 2). When “Not tested” and “Tested positive” were combined to compare among the three types of chemical senses, we found that the occurrences of chemosensory dysfunction of smell and taste were not largely different from each other, and dysfunction in chemesthesis was reported significantly less than expected values. The differences in the occurrences of chemosensory dysfunction were statistically significant (Chi-square test, χ2=390.616, df=4, P<0.001). Thus, the sense of smell and taste are more vulnerable to COVID-19 compared to chemesthesis.

**Fig. 2.**
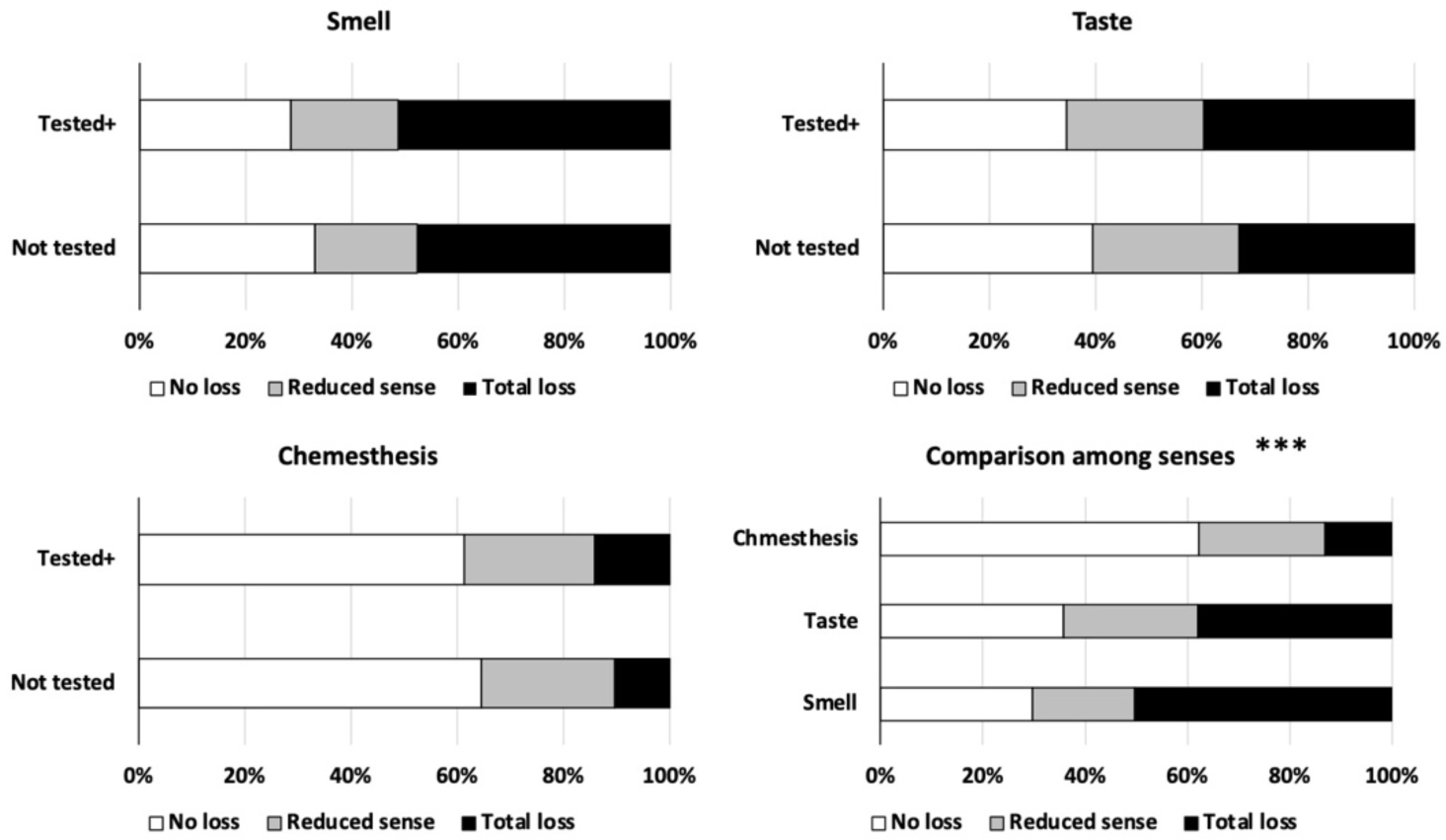
Self-reported senses of smell, taste, and chemesthesis by the “Not tested” and “Tested positive” groups and comparison of the occurrences of chemosensory dysfunction in the sense of smell, taste, and chemesthesis. ***: Chi-square test, P<0.001

#### v) Intake of foods and beverages

Based on the goal of this survey to determine whether there are differences in the consumption of each type of beverages and food materials, we focused on the differences between the “Tested positive” group and others, especially with “Did not get COVID” but also with “Not tested” group since the symptoms were lighter in this group compared to the “Tested positive” group.

#### a) Beverages

Figure 3 shows the results on beverage consumption. Other than hot chocolate, we found significant differences among the 3 groups of participants (Chi-square tests, tea, χ²=54.528, df=6, P<0.001; herbal tea, χ²=31.060, df=6, P<0.001; coffee, χ²=22.207, df=6, P=0.001; cider/apple cider, χ²=79.553, df=6, P<0.001, alcohol, χ²=29.920, df=6, P<0.001; hot chocolate, χ²=9.771, df=6, N.S.) (Supplementary Table 3). “Tea” and “coffee” are, in general, consumed daily more than other types of beverages.

**Fig. 3.**
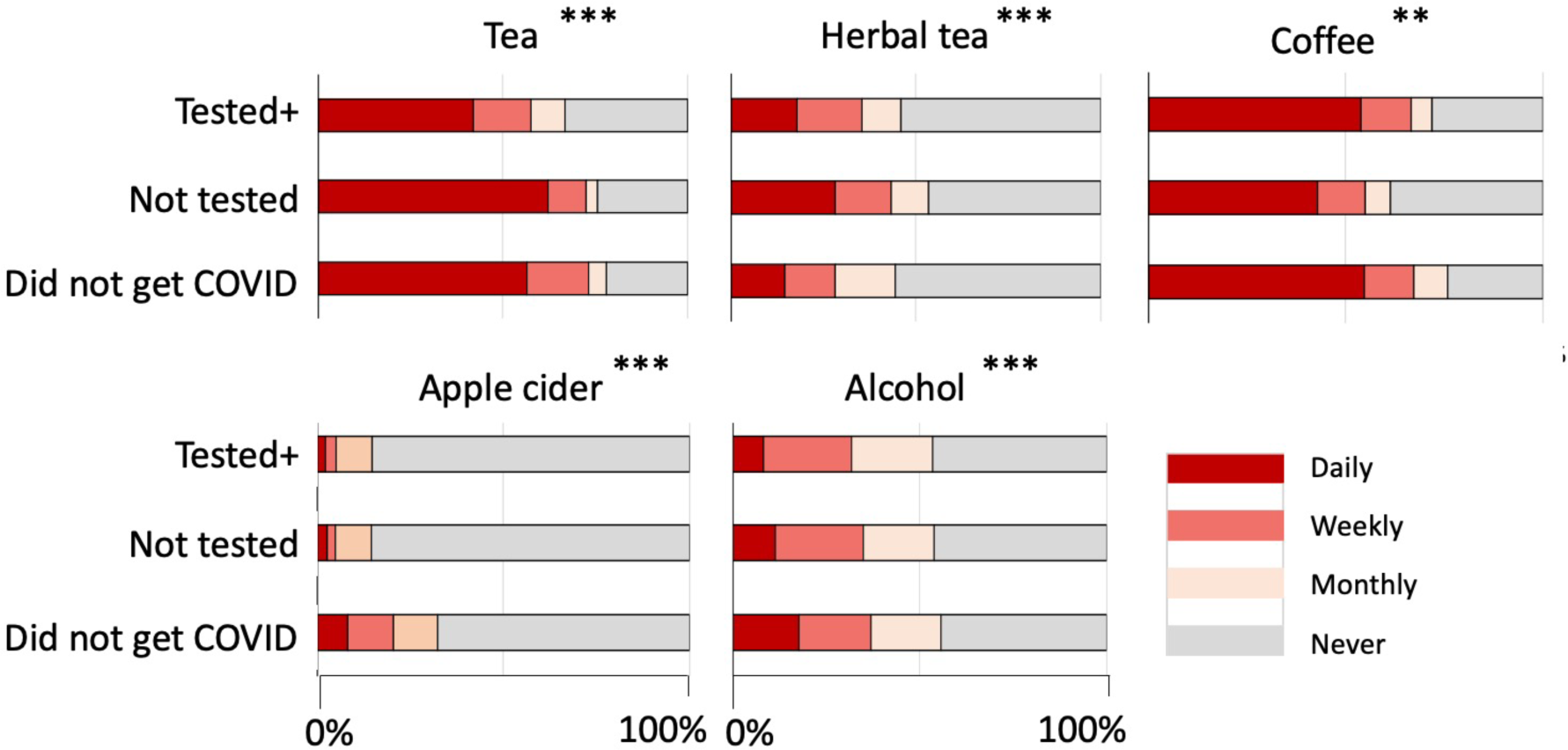
Intake of tea, herbal tea, coffee, apple cider, alcohol, and hot chocolate. **: P<0.01, ***: P<0.001

Table 1 shows the Chi-square scores of each cell in the Chi-square tests and the color markers show whether the observed values were higher or lower than the expected values, contributing to the statistical significance in the Chi-square tests. The darkness of the color was arbitrarily classified in three levels where higher Chi-square scores were assigned darker color to show larger discrepancy from the expected values. “Tea” was significantly more daily consumed by participants who “Did not get COVID” and “Not tested” (Figure 3 and Table 1). Although herbal tea is often discussed with its beneficial effects on health, herbal tea was significantly less consumed by the “Did not get COVID” group (Figure 3 and Table 1). Although the overall consumption was much less than other beverages, apple cider was found to be significantly more consumed by “Did not get COVID” group participants. Coffee did not show specific tendencies among groups. Daily consumption of alcohol reported was higher in the “Did not get COVID” group. Hot chocolate was not consumed so frequently by any of the groups.

**Table 1.**
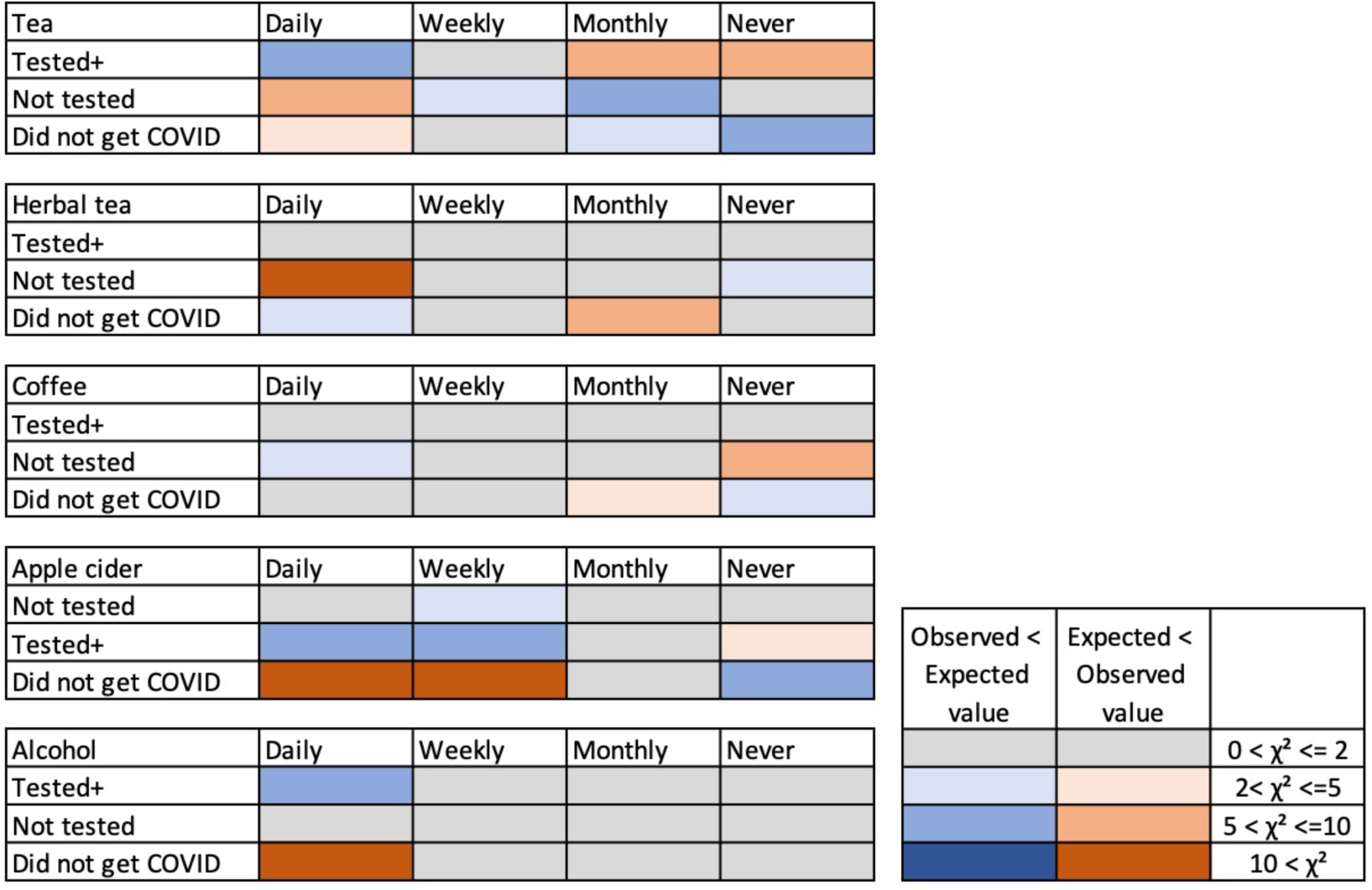
Results on Beverages. Discrepancy of observed values from expected values in the chi-square test. The colors indicate that the observed values were higher than the expected values (smaller to larger discrepancy indicated by pink to red color) and the observed values lower than expected values (smaller to larger discrepancy indicated by light to dark blue). Grey color indicates there were no or negligible discrepancies.

#### b) Foods

There was an overall tendency that the “Did not get COVID” group daily consumed leaf, root, other vegetables, beans and peas, nuts and seeds, spices and herbs, other foods, such as mushrooms, and fermented food significantly more than the expected values compared to other two groups (Chi-square test, leaf vegetable, χ²=23.957, df=6, P<0.001; root vegetable, χ²=15.828, df=6, P=0.015; other vegetable, χ²=13.848, df=6, P<0.05; beans and peas, χ²=47.242, df=6, P<0.001; nuts and seeds, χ²=10.160, df=6, N.S.; herbs and spices, χ²=36.757, df=6, P<0.001; cereals and grains including barley, χ²=21.060, df=6, P=0.0018; other foods, χ²=51.960, df=6, P<0.001; fermented foods, χ²=62.945, df=6, P<0.001) (Figure 4 and Supplementary Table 2). Differences from the expected values showing that the “Did not get COVID” group daily consumed more compared to other two groups were especially large in leaf vegetables, beans and peas, herbs and spices, other foods, and fermented foods (Table 2).

**Fig. 4.**
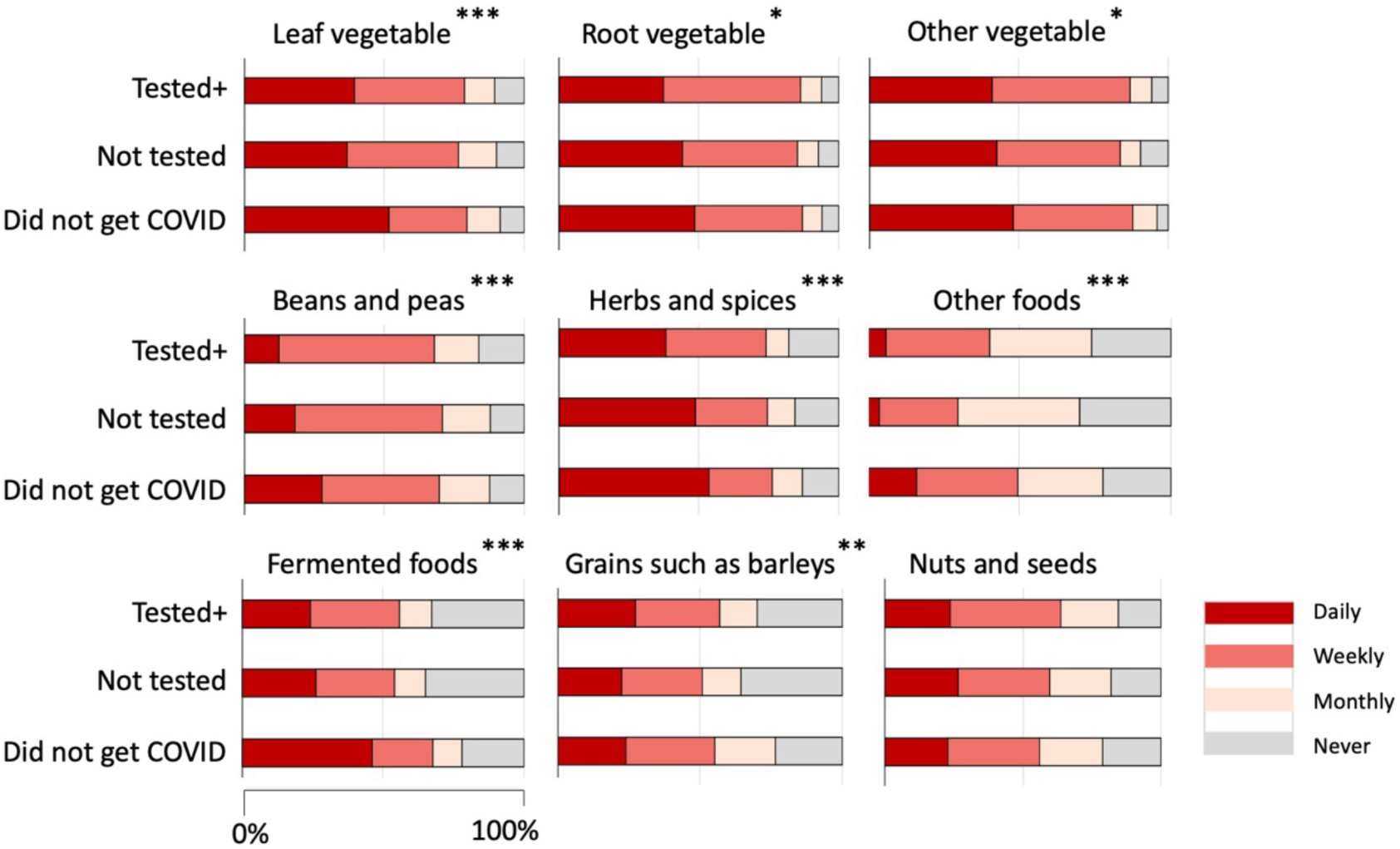
Frequency of consuming various types of foods by the three groups. *: P<0.05, **: P<0.01, ***: P<0.001.

**Table 2.**
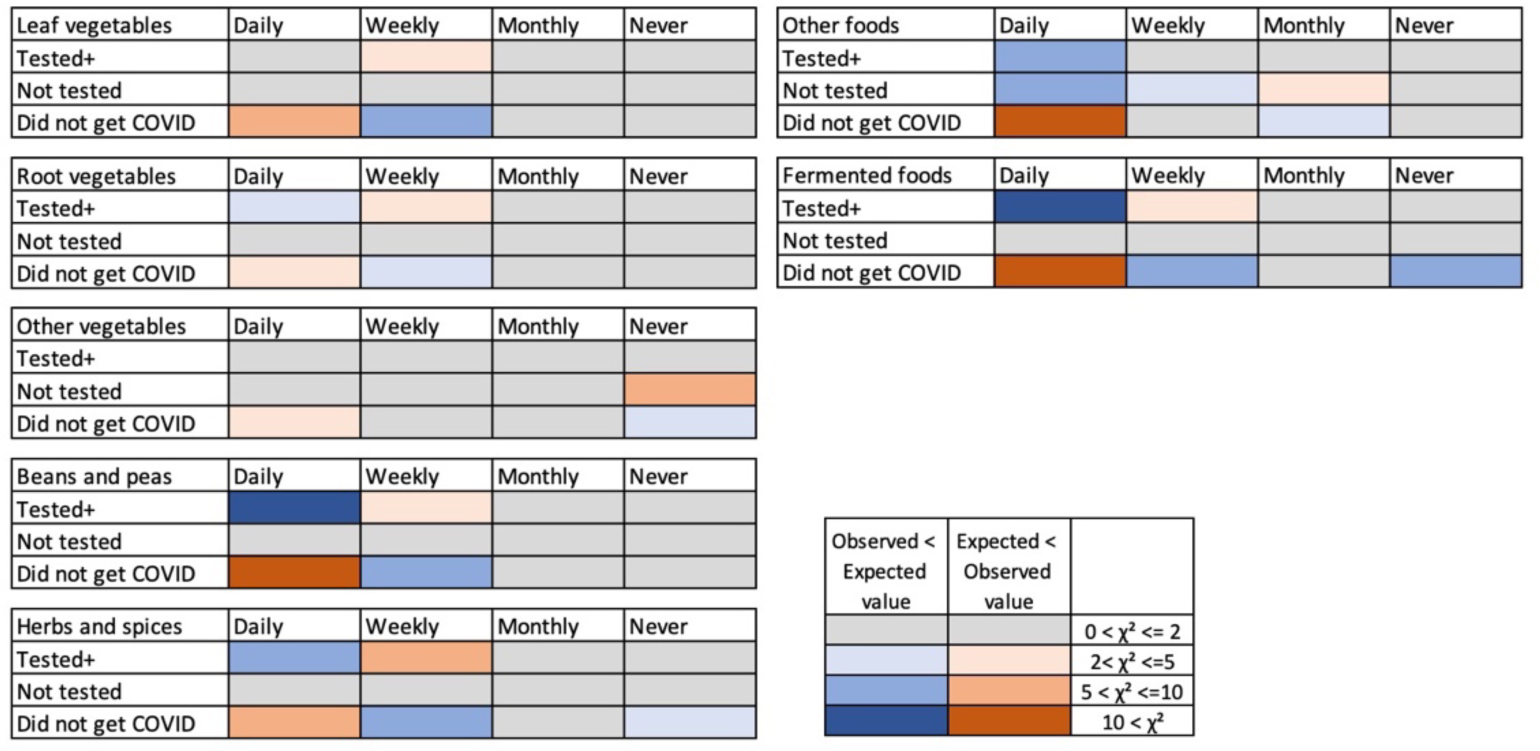
Results on Foods. Discrepancy from expected values. The colors indicate that the observed values were higher than the expected values (smaller to larger discrepancy indicated by pink to red color) and the observed values lower than the expected values (smaller to larger discrepancy indicated by light to dark blue). Grey color indicates there were no or negligible discrepancies.

In fruits, the consumption of citrus fruits, berries, pome fruits, stone fruits, and other fruits showed statistically significant differences in the frequency of eating among the health conditions of the participants (Chi-square test, Citrus fruits, χ²=31.541, df=6, P<0.001; berries, χ²=21.167, df=6, P=0.0017; pome fruits, χ²=25.777, df=6, P<0.001; stone fruits, χ²=8.617, df=6, N.S., tropical fruits, χ²=8.001, df=6, N.S.; other fruits, χ²=16.516, df=6, P<0.05) (Figure 5, Supplementary Table 3). Comparison of the observed values and the expected values (chi-square scores), however, did not indicate that the “Did not get COVID” group daily ate fruits more than other groups (Table 3). Other than the stone fruits, the “Did not get COVID” group ate fruits rather less than other two groups (Table 3).

**Fig. 5.**
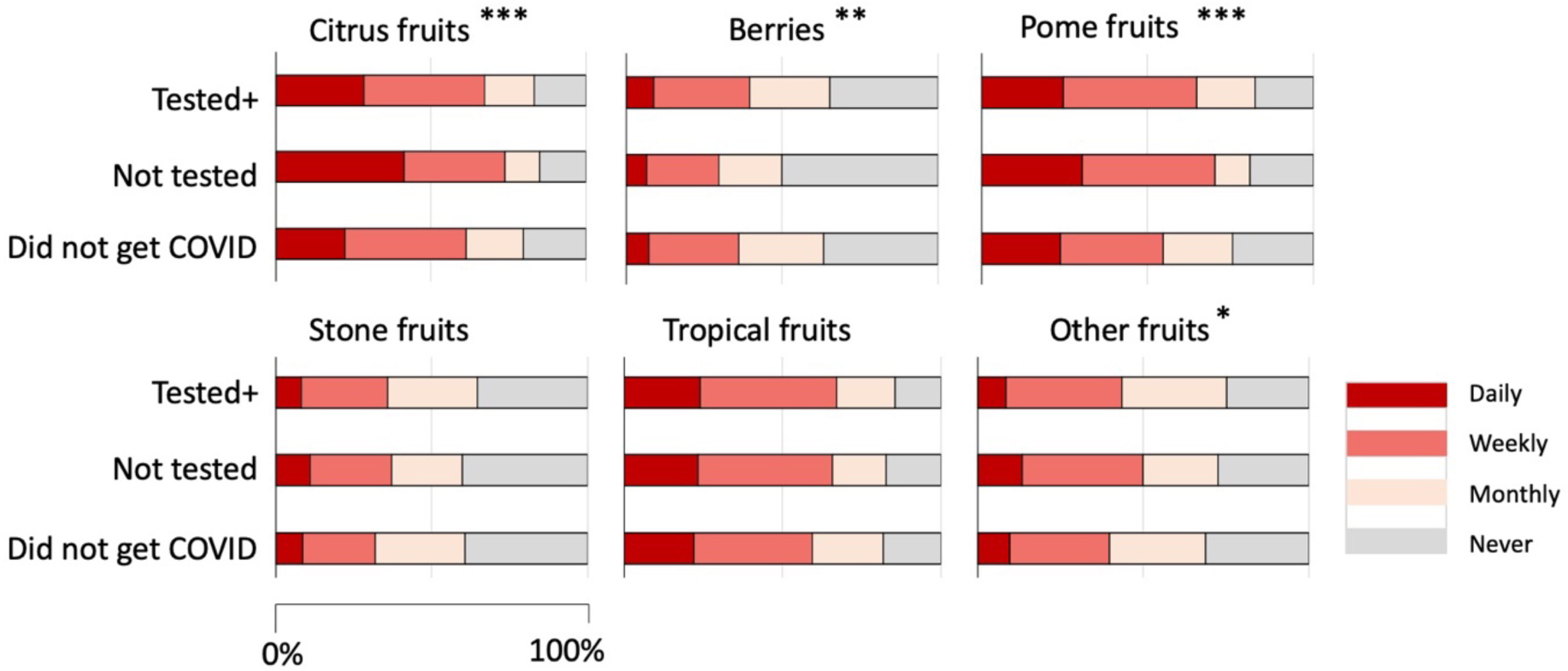
Frequency of consuming fruits by the three groups. *: P<0.05, **: P<0.01, ***: P<0.001.

**Table 3.**
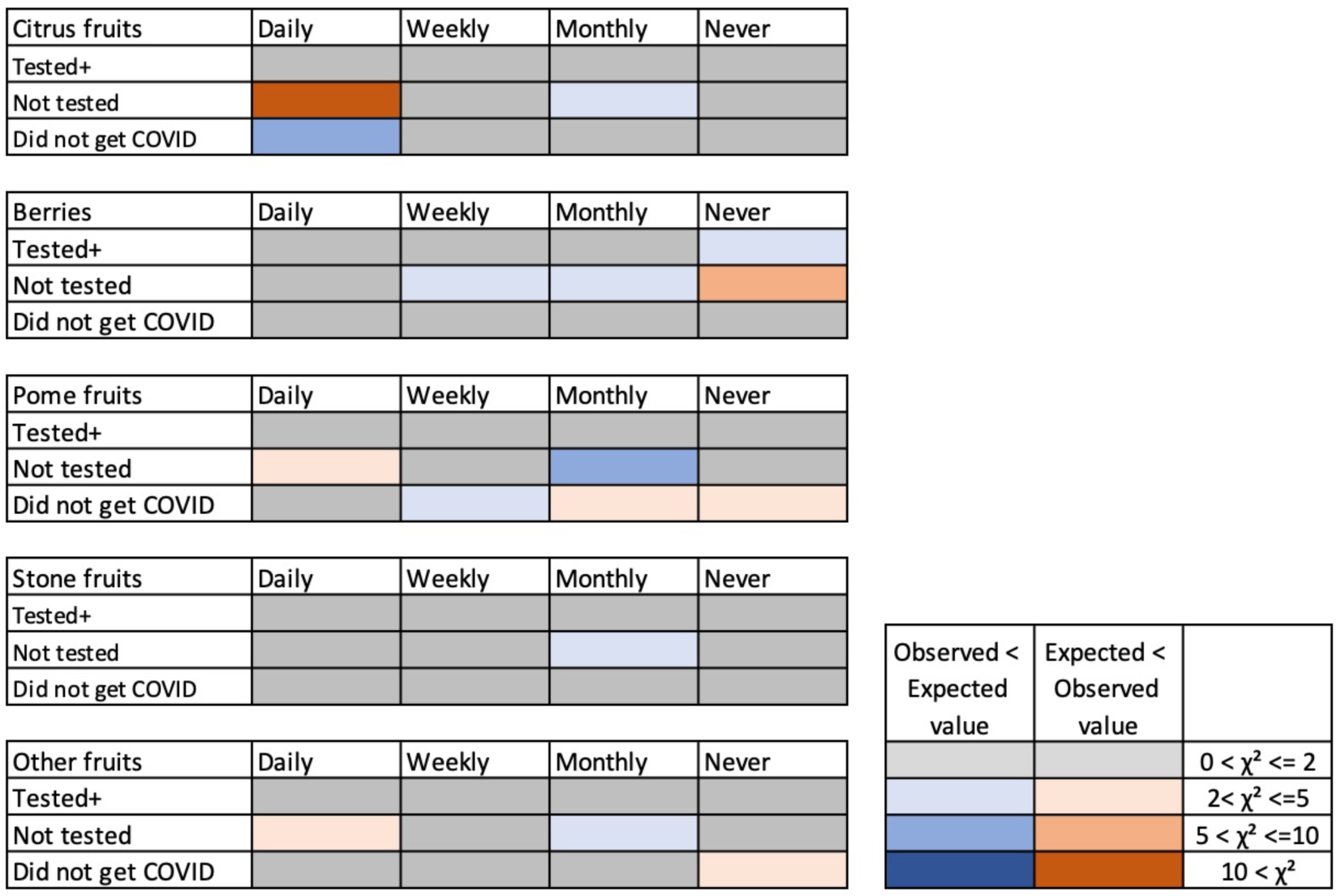
Results on Fruits. Discrepancy from expected values. The colors indicate that the observed values were higher than the expected values (smaller to larger discrepancy indicated by pink to red color) and the observed values were lower than the expected values (smaller to larger discrepancy indicated by light to dark blue). Grey color indicates there were no or negligible discrepancies.

#### c) Herbal medicines, supplements, and essential oils

There were significant differences in the way that herbal medicines and supplements were taken, and essential oils were used by the three groups (Chi-square test, herbal medicine, χ²=18.872, df=6, P=0.004; supplements, χ²=38.246, df=6, P<0.001; essential oils for prevention, χ²=10.904, df=6, N.S.; essential oils for recovery, χ²=4.807, df=3, N.S.) (Supplementary Figure 2 and 3, Supplementary Table 3). Comparison of the Chi-square scores revealed that the “Did not get COVID” group took herbal medicines and supplements or used essential oils significantly less than the expected values (Supplementary Table 4).

### Comparison of countries

Next, we conducted analyses of the results for all the versions of languages separately. This was based on our interest in the cultural differences in the eating and drinking habits and their possible relationship with the infection rate in various countries, which became often discussed on social media at the beginning of the pandemic. The nationality of the participants of each language version of the survey did not necessarily mean that they were ethnically affiliated to a certain country (for example, Spanish is spoken in many countries). We determined the ethnicity of the participants in each language version of the survey using the answers to the questions on ethnicity and residing location. We found that, other than the English version, over 90% of the participants of each language version of the survey were ethnically affiliated to one country (Supplementary Table 5). Although Spanish is spoken in many countries, the participants in this survey were over 90% affiliated to Spain. The participants of the Persian version were also over 90% affiliated to Iran. This suggests that it is possible to expect that the results of each language version, other than the English version, may be able to represent geographical and cultural differences in the eating and drinking habits. The participants of the English version reported their ethnicity as Greek, Asian, Indian, Vietnamese, Israeli, Irish/German, Bosnian, and Montenegrin (Supplementary Table 5).

As the number of participants for some of the language versions of the survey was too few to analyze the data, we focused on the results of the language version that had more than 100 participants and excluding the English version because of the mixed ethnicity nature. Thus, the comparison of countries was conducted using the data of the Indian English, Italian, Japanese, Persian, Russian, and Spanish versions of the survey to represent the six countries of India, Italy, Japan, Iran, Russia, and Spain.

#### a) Beverages

We focused on the percentage of participants who reported that they have been drinking the type of beverage daily. We could see a large difference among the countries in the results. Figure 6 shows the results of tea and coffee. In Japan, the percentage of participants who reported to drink tea or coffee daily was similarly high. India, Russia and Iran showed a high percentage of drinking tea daily but much less percentage of drinking coffee daily. Contrarily, in Italy and Spain, the percentage of drinking coffee daily was much higher than that of tea. Although the cultural differences were clear and suggested there might be differences in the tendencies of consumption if we take these differences into consideration, the percentages of daily consumption of tea and coffee among the health conditions did not show specific tendencies that suggest the benefits of tea or coffee to prevent contracting COVID-19.

**Fig. 6.**
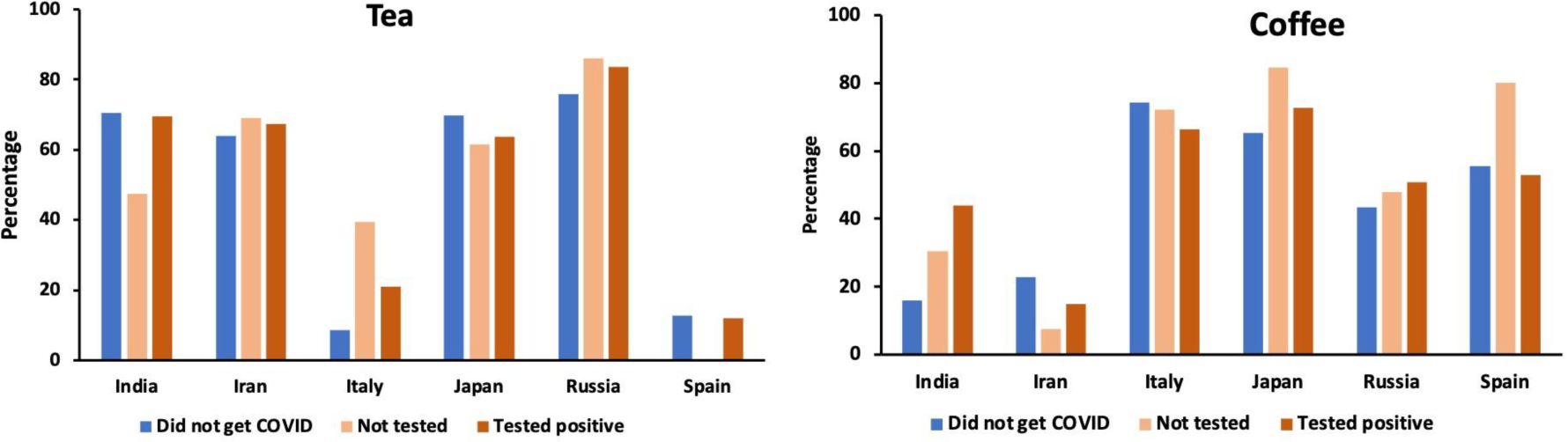
Percentage of participants in each group who have been drinking tea or coffee daily.

Table 4 shows the magnification of the discrepancy between the expected values and observed values (Chi-square scores) of the “Did not get COVID” group of the six countries. Japan and Russia showed higher than expected values in the answers of “Daily” in tea, whereas Italy and Japan showed higher than expected values in the answers of “Daily” in coffee. The observed value of “Daily” in coffee by participants in Spain was not higher than the expected values but the observed value of “Daily” in tea was much lower than the expected value in Spain, suggesting that tea is not favored in Spain. These results also indicate the drinking habits of both tea and coffee in Japan, drinking coffee in Italy, and drinking tea in Russia.

**Table 4.**
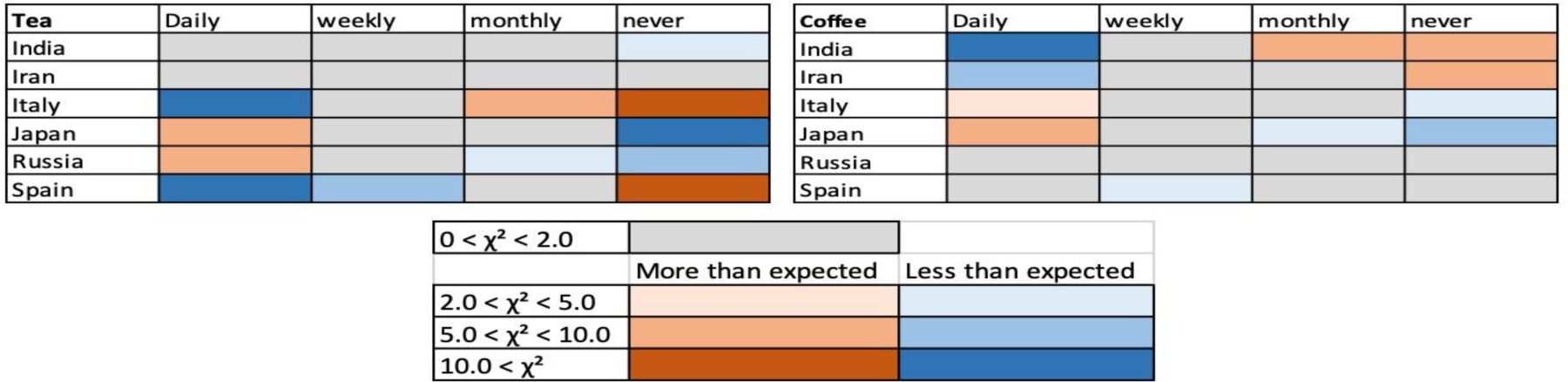
Results on Tea and coffee. Discrepancy from expected values - comparison among the “Did not get COVID” group of each country. The colors indicate where the observed values were higher than the expected values (smaller to larger discrepancy indicated by pink to red color) and where the observed values were lower than the expected values (smaller to larger discrepancy indicated by light to dark blue). Grey color indicates there were no or negligible discrepancies.

#### b) Foods

In the overall section, the results of leaf vegetables, root vegetables, fermented foods, beans and peas, cereals and grains including barley, and herbs and spices showed higher percentages of daily consumption by the “Did not get COVID” group. When the results were split among countries, we found large differences in the tendencies among them (Chi-square test, leaf vegetables, χ²=138.382, df=15, P<0.001; root vegetables, χ²=74.398, df=15, P<0.001; fermented foods,. χ²=129.664, df=15, P<0.001; beans and peas, χ²=121.634, df=15, P<0.001; herbs and spices, χ²=110.345, df=15, P<0.001; cereals and grains including barley, χ²=73.772, df=15, P<0.001; other vegetables, χ²=51.673, df=15, P<0.001). Overall, Japan showed higher intake of all the categories of foods other than cereals and grains including barley, compared with other countries but there were no statistically significant differences among the health conditions other than the results on herbs and spices (Figure 7 and Supplementary Figure 3 for the rest of the categories). The results by the “Did not get COVID” group of Italy showed clearly higher percentages of daily consumption in the 7 categories of foods although the overall percentages were smaller than the results in Japan (Figure 7).

**Fig. 7.**
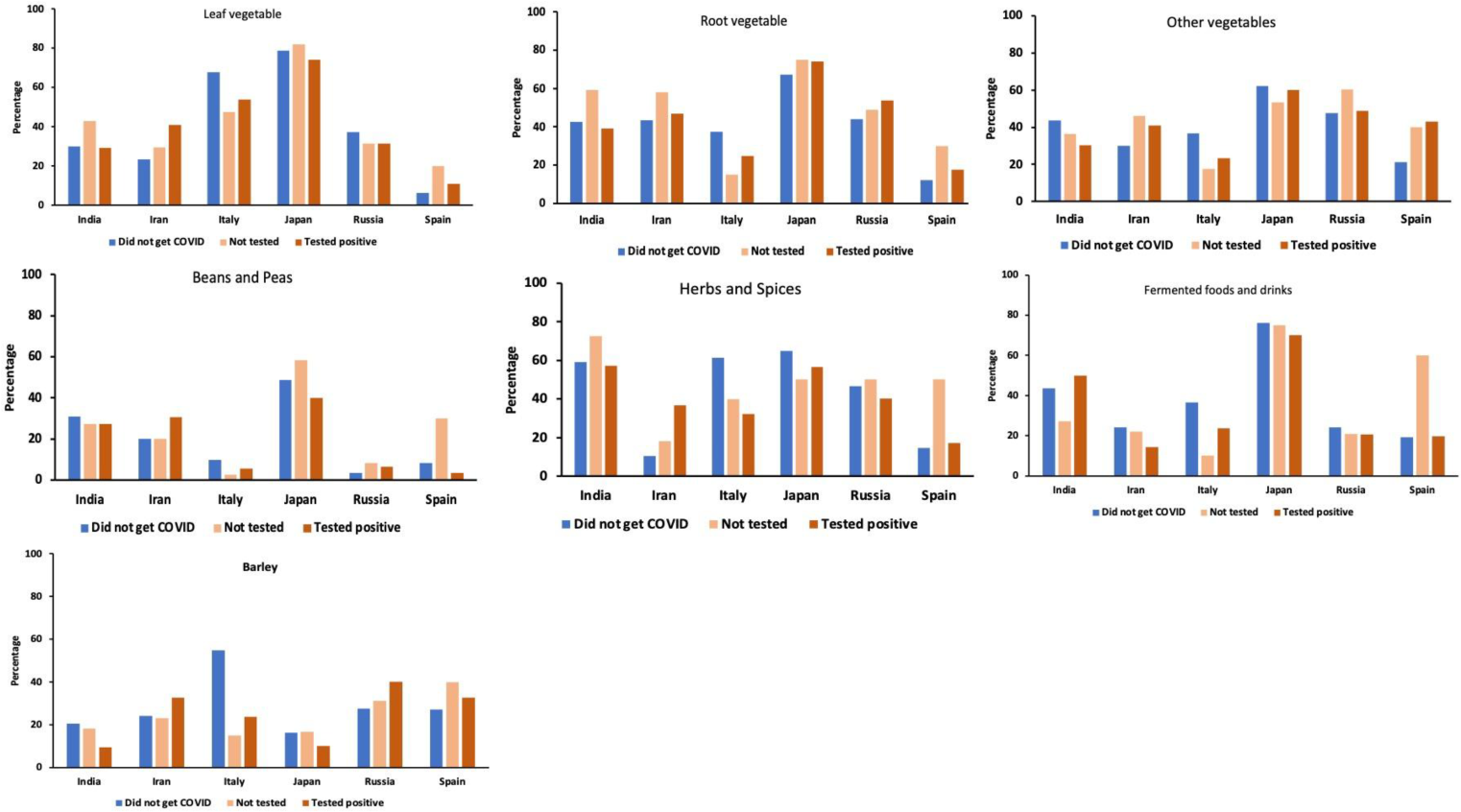
Percentage of participants who were daily eating leaf vegetables, root vegetables, fermented foods, herbs and spices, beans and peas, and other foods such as mushrooms and olives.

Table 5 shows the discrepancy between the expected values and observed values in the answers by the “Did not get COVID” group of the 6 countries. The “Did not get COVID” group of Japan showed higher daily consumption of leafy vegetables, root vegetables, other vegetables, beans and peas, herbs and spices, and fermented foods, and Italy showed a higher consumption of cereals and grains including barley (Table 5). The “Did not get COVID” group of Spain showed a lower daily consumption of leaf vegetables, root vegetables, other vegetables, beans and peas, herbs and spices, and fermented foods, showing a clear contrast to the results of the “Did not get COVID” group of Japan.

**Table 5.**
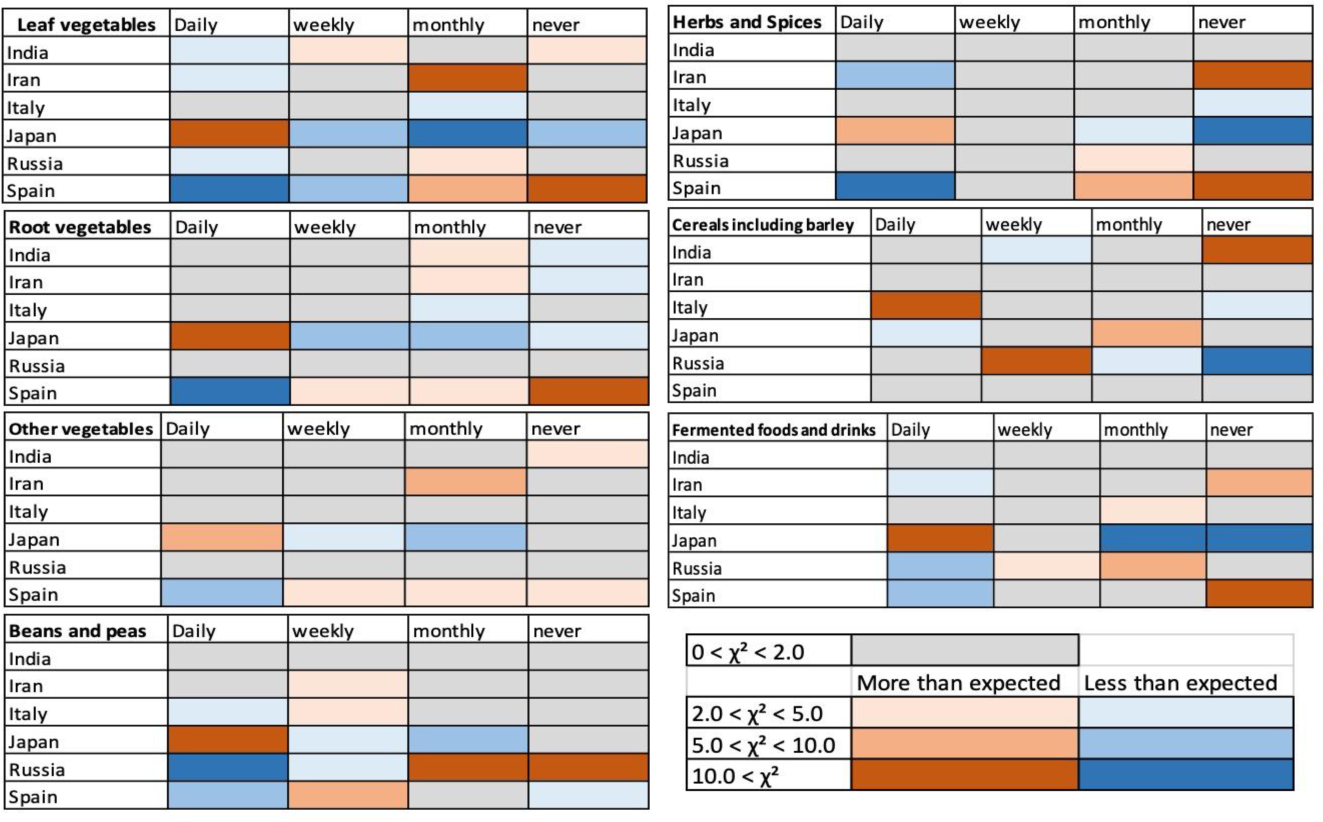
Results on Foods. Discrepancy from expected values - comparison among the “Did not get COVID” group of the six countries. The colors indicate where the observed values were higher than the expected values (smaller to larger discrepancy indicated by pink to red color) and where the observed values were lower than the expected values (smaller to larger discrepancy indicated by light to dark blue). Grey color indicates there were no or only negligible discrepancies.

Such differences among countries also suggest the possibility that, in the results where we did not see clear differences among health conditions in the overall analysis section above, the differences depending on the country might have compensated each other, and produced that lack of tendencies. Indeed, there were large differences in the percentages of participants who ate the food types daily that did not show specific tendencies in the overall section depending on countries and health conditions (Supplementary Figure 3). Interestingly, the results of the survey in Japanese showed low percentages of daily consumption of all the types of fruits regardless of the health condition type, compared to other countries (Supplementary Figure 3). Considering the high percentage of daily consumption in all types of vegetables in the Japanese version, “daily consumption of vegetables and less frequent consumption of fruits” seemed to be the characteristics of food consumption in Japan. The results of the Italian version of the survey showed higher daily consumption of tropical fruits, barleys, other fruits, and other vegetables by the “Did not get COVID” group than other two groups (Supplementary Figure 3). These results suggested striking differences in the eating habits of the “Did not get COVID” groups in Italy and Japan.

#### c) Herbal medicines, supplements, and essential oils

Herbal medicines have been used for centuries in countries all over the world. In the questions about the usage of herbal medicines, we found that the daily usage is rather high in India and Iran (Supplementary Figure 4). “Not tested” group participants in Iran also showed high daily usage of herbal medicine. Daily usage of supplements had the tendency to be higher in the “Not tested” and “Tested positive” groups than the “Did not get COVID” group, and low in the Spanish version (Supplementary Figure 4). Daily usage of essential oil for prevention and recovery was low overall with the tendency to be higher in the “Tested positive” group (Supplementary Figure 4).

### Comparison with recovery

There were significant differences in the self-reported days for recovery among the countries (Kruskal Wallis test, “Not tested”, H=20.397, df=5, P=0.001; “Tested positive”, H=21.808, df=5, P=0.001; results of pairwise comparison as well as medians of the days to recover are shown in Figure 8). This time in days for recovery was not necessarily the time until the PCR tests turned negative, but it was the self-reported time length until recovery. The recovery time reported by participants of both “Not tested” and “Tested positive” groups in Japan was statistically significantly shorter than in countries other than India (Figure 8). The recovery time of the “Not tested” group of India was statistically shorter than all other countries except Japan, and the recovery time of the “Tested positive” group of Iran was significantly longer than other countries although the median was not different (Figure 8). The median recovery time of the “Not tested” group of Spain was long, but it was not statistically significantly different from the recovery time of the “Not tested” group of Italy and Russia (Figure 8).

**Fig. 8.**
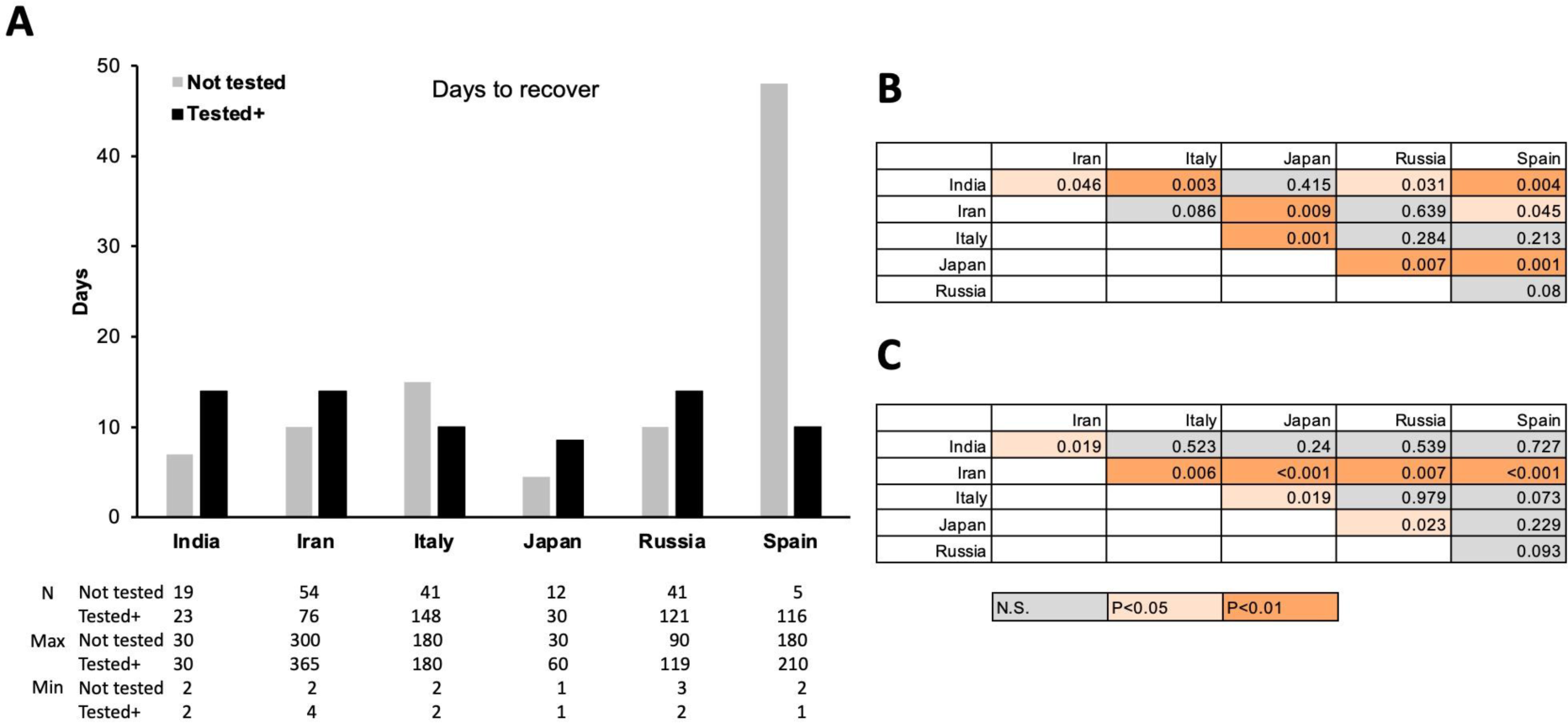
Median of the self-reported time (days) until recovery of “Not tested” and “Tested positive” group participants (a) and tables showing statistical differences of pairwise comparisons of the “Not tested” (b) and “Tested positive” (c) groups.

Other than the results of Spain, the median of the number of days until recovery in the “Tested positive” group ranged from 8.5 days (Japan) to 14 days (India, Iran, Russia), and those in the “Not tested” group ranged from 4.5 days (Japan) to 15 days (Italy). The minimum of the reported days ranged from 1 to 4 and the maximum of the reported days ranged from 60 days to 365 (one year), excluding the participants who reported they are still not recovered, most likely with Long-COVID. The days until recovery between the “Not tested” and “Tested positive” groups were significantly longer in the “Tested positive” group than the “Not tested” group in India (Mann-Whitney U-test, P=0.014) and Iran (Mann-Whitney U-test, P<0.001), and not significantly different in other countries. Overall, these results indicate the significant differences among countries and significantly short time to recover in Japan and India, and longer time to recover by the “Tested positive” group in India and Iran.

These differences in the time to recover among countries could be due to the type of the virus (original and variants that emerged later) they contracted and the availability of the medicines especially at the early stage of the pandemic. Figure 9 shows the numbers of answers to the question asking when the participants (“Not tested” and “Tested positive” group combined, and English version excluded as the location of the participants is not clear) contracted COVID-19 (Figure 9). Large number of participants in Italy contracted COVID-19 early in 2020 whereas the participants from India, Iran, and Japan show a high peak in ‘after December’ 2021. Spain also showed a high peak in spring 2020 and a smaller peak in ‘after December 2021’. The number of the participants from Russia shows multiple peaks with slightly higher numbers in spring 2020 and after December 2021.

**Fig. 9.**
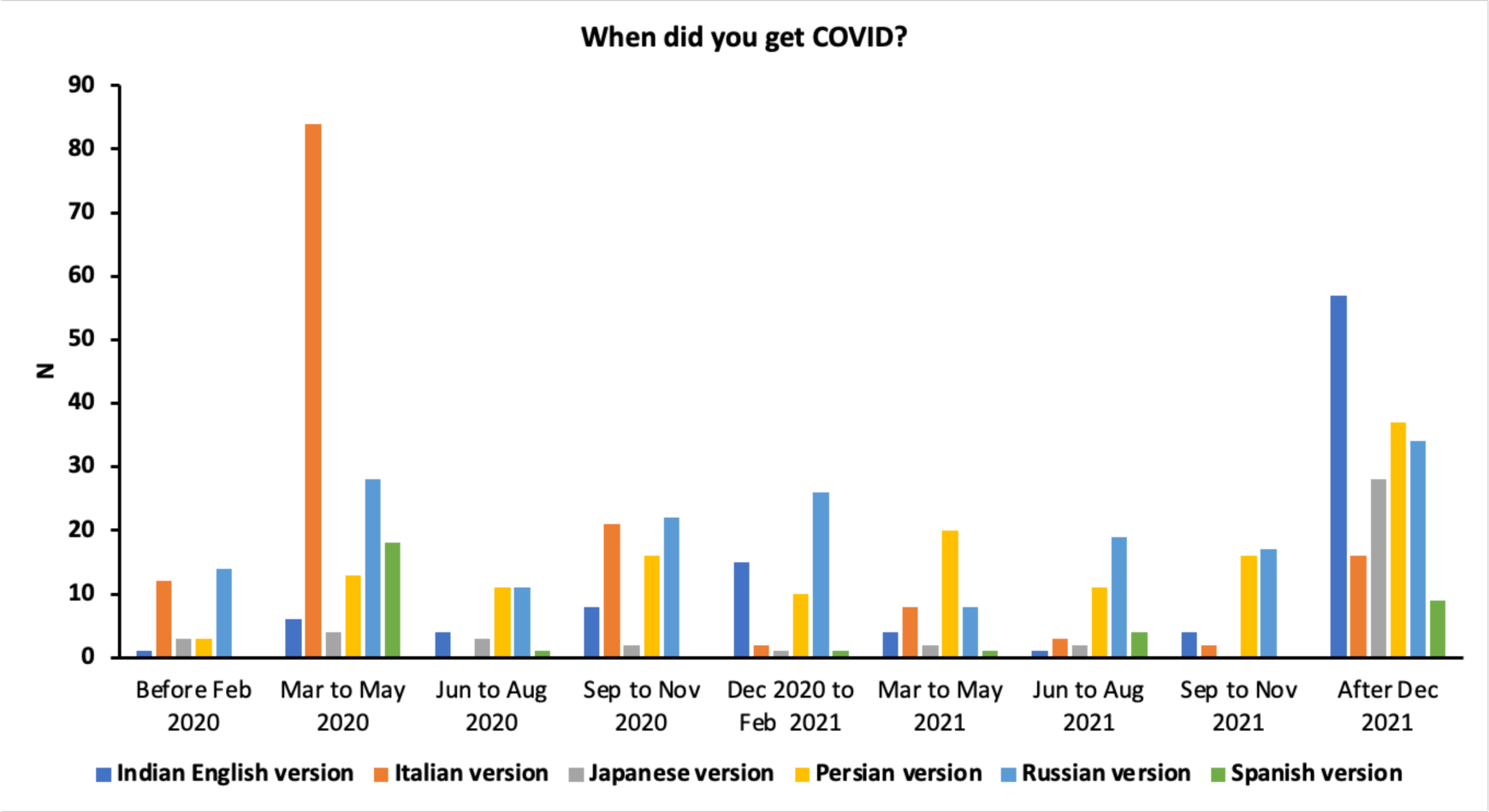
Reports on the dates the participants contracted COVID-19 (“Not tested” and “Tested positive” group combined).

Although the timing of contracting COVID-19 can affect the time that it may take to recover, there is also a possibility that daily consumption of certain types of foods and beverages can have effects on facilitating recovery from COVID-19. And, if so, there is a possibility that the more people take these foods, the time to recover may become shorter. This could be especially important in case of the “Not tested” group as they were most likely not prescribed medicines, because they were not tested. The food and drink categories that are more frequently consumed in Japan and/or India could suggest the types of foods and beverages that may have such possibly facilitating effects. Figure 10 shows the food that were taken daily and had higher percentages of daily consumption in India and Japan. Beverages did not show specifically higher percentages in Japan and in India than other countries. Hence, they are not shown in Figure 10. Herbs and spices and fermented food/beverages were consumed daily at higher percentages in India and in Japan compared to other countries (Figure 10a). Figure 10b shows the categories of foods that were especially high in Japan and Figure 10c shows the categories of foods that were higher in India than other countries, although the percentages were not extremely high, compared to other food categories. These categories of foods may have had the effects on preventing as well as facilitating recovery from COVID-19 and that these could be the candidate foods for further tests in future.

**Fig. 10.**
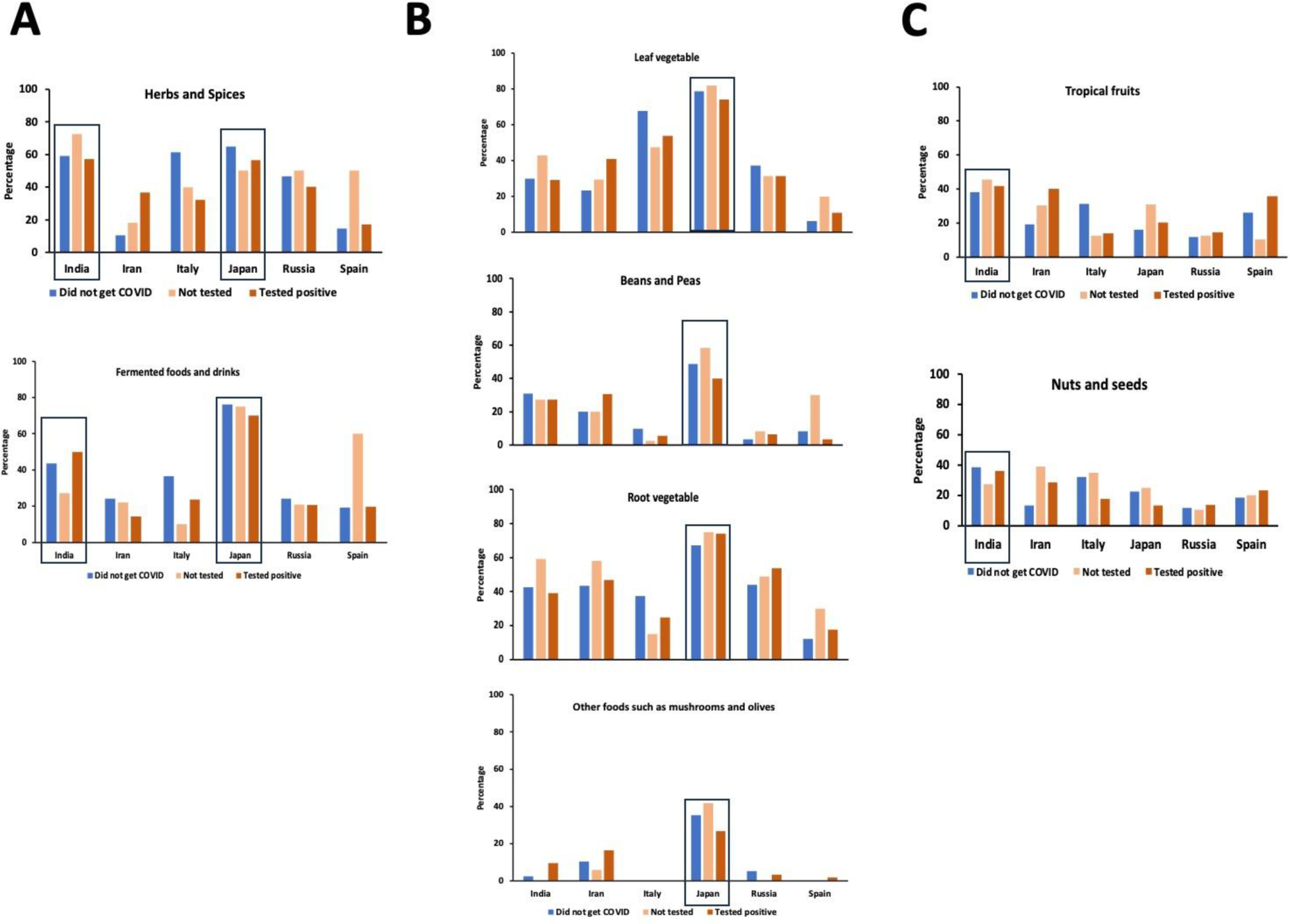
Daily consumption of foods in each country. (a) Food categories that showed high percentages of daily consumption in India and Japan. (b) Food categories that showed high percentages of daily consumption in Japan than other countries. (c) Food categories that showed higher percentages of daily consumption in India than other countries. India and Japan are highlighted by squares.

Dysfunction in the chemical senses is now well-known as one of the major symptoms of COVID-19. Large number of the participants from Italy replied to have experienced chemosensory dysfunction, and participants from Russia and Spain also replied with a high number of chemosensory dysfunctions. These are the countries where many of the participants reported they contracted COVID-19 early in 2020, when the pandemic was more virulent and the influences of contracting the virus were strongest, and this could be the reason for the higher incidences of chemosensory dysfunction (Supplementary Figure 5).

## Discussion

The key findings from our survey were as follows: 1) higher percentage of people who did not get COVID-19 daily ate various types of vegetables and fermented foods/beverages, 2) in India and Japan, the people who did not get tested and who tested positive showed significantly shorter time to recover from COVID-19, 3) the people who tested positive or did not get tested and recovered rather quickly were eating/drinking these vegetables and fermented foods/beverages significantly more daily, 4) fruits are known to have beneficial effects on health, but they were not specifically consumed more by those who did not get COVID-19, 5) teas are also known to have beneficial effects on health and on COVID-19, and people who did not get tested and who did not get COVID-19 were found to have a higher daily intake of tea, 6) people who tested positive reported more severe symptoms compared to people who did not get tested, and 7) there were significant differences in the number of days to recover among countries in both the group that did not get tested group and the group that tested positive.

Tea and coffee were found to be the most preferred beverages to drink daily, although there were some country-dependent differences about which is more preferred. In our survey, we found that tea was significantly more consumed daily by the people who did not get COVID-19 and those who were not tested than by the participants who tested positive to COVID-19. Although it is difficult to conclude from survey data whether it had some effects or not, there were significant differences in the severity of the symptoms between people who tested positive and those who did not get tested. Previous studies have shown that (-) epigallocatechin gallate (EGCG), theasinensin A (TSA), and galloylated theaflavins such as theaflavin 3,3’-di-*O*-gallate (TFDG) included in teas have anti-SARS-CoV-2 activities [10, 11, 21, 42, 43]. The differences in the processing methods generate large differences in the concentration of chemical composition in green tea, yellow tea, white tea, oolong tea, and black tea [44]. TSA and TFDG are catechin derivatives that increase by enzymatic oxidation during the processing of the tea leaves, and thus EGCG is decreased in black tea [44, 45]. The studies showing that EGCG, TSA, and TFDG have anti-SARS-CoV-2 effects indicate that these changes in the amount included by differences in the processing methods do not affect the potential positive effects of tea. Importantly, it should be noted that there are studies showing that milk casein blocks the anti-SARS-CoV-2 effects of tea because of the binding of them to casein [46]. These studies suggest the possible effects of teas on preventing and suppressing severity of COVID-19, but also indicate that depending on how the tea is prepared (for example, milk tea and chai tea, which adds milk), the effects can be blocked.

Although in our survey the results on coffee did not show its effects on facilitating recovery from COVID-19 or suppressing infection, a study using a large amount of data (n>37K) in the UK Biobank on dietary habits and COVID-19 infection rate has shown a significant negative correlation between the drinking habit of coffee and the infection rate, *i.e*., people drinking coffee were less infected to COVID19 [47]. An *in vitro* study has also shown that coffee, caffeine, or diluted human serum samples collected from healthy adults who drank coffee suppressed infection of SARS-CoV-2 [22]. It could be that the lack of tendency in our project is due to the smaller data size, and it is also possible that the differences are due to the international nature of our study, or possible interactions with other beverages or foods or the way of preparation like in case of teas.

An important result in this study is that the people who did not get COVID-19 showed general trend to consume leaf, root and other vegetables, mushrooms, fermented foods/beverages, beans and peas, and herbs and spices daily and that the people who tested positive and those who did not get tested and consumed them daily recovered faster. This tendency of inverse relationship between intake of vegetables and COVID-19 was also found in previous COVID-19 literature. Tadbir Vajargah et al (2022) [48] found that the severity of COVID-19 symptoms of 250 hospitalized patients negatively correlated with the amount of consumption of vegetables as well as fruits and dietary fiber. They also recovered faster, leading to shorter hospitalization and convalescence periods. A meta-analysis study, which was conducted before the COVID-19-pandemic, has also shown that higher intakes of fruits and vegetables reduce proinflammatory cytokines, and improved the immune system profiles [49]. In a survey-based study focusing on six European countries, researchers found that people who were on plant-based diet or pescatarian diet (plant-based diet with fish) had milder symptoms of COVID-19 [12]. The results of our study are in line with these studies and suggest the importance of vegetable intake and also highlight the importance of studies on the key phytochemicals or combinations of phytochemicals that contributed to these differences.

In case of fermented foods and beverages, there are additional factors that are necessary to take into consideration, *i.e*., the roles of bacteria. There are also animal-based fermented foods/beverages, which may have different effects compared to those that are plant-based. Muhialdin et al. (2021) [35] summarized the antiviral activities of probiotic bacteria in fermented foods. They suggested that the antiviral effects of fermented foods could be exerted directly through the viral neutralization by the bioactive compounds and indirectly through improving the innate immune system [35]. It should be noted that the fermented foods/beverages category was one of the two most commonly reported categories in India and Japan; the two countries that showed significantly faster recovery from COVID-19. This suggests the importance of further studies on fermented foods/beverages.

Considering the well-known beneficial effects of fruits on health, we hypothesized that we would find higher daily consumption of fruits, but, interestingly, with the exception of stone fruits, daily fruit consumption was rather low compared to vegetables and fermented foods. In fact, there are studies that found the negative association of fruits consumption and COVID-19 [48]. In the study that identified foods that contain chemical compounds with bioactive properties that resemble clinically approved drugs for COVID-19, they identified 52 phytochemical compounds with anti-SARS-CoV-2 effects [18]. They reported that flavonoids, coumarins, stilbenes, indoles, and phenolic acids are the major groups of phytochemicals with anti-COVID-19 effects. Examples they listed are quercetin, kaempferol, myricetin of flavonols, luteolin and apigenin of flavones, procyanidin B2 of flavanols, naringin of flavanones, daidzein, genistein, legumelin of isoflavonoids, trans-resveratrol of stilbenes, 3-indolecarbinol of indoles, and gallic acid of phenolic acids. Many of these phytochemicals with bioactive properties against SARS-CoV-2 are included in fruits (berries, citrus fruits, pome fruits, stone fruits, and tropical fruits). It could be that, similarly to coffee, there may be some differences in the images of fruits among countries (for example, fruits are called ‘mizu-gashi’ in old Japanese language, which means ‘watery sweets/desserts’). Japanese participants reported very high daily intake of all types of vegetables and fermented foods/drinks in comparison to other countries, but daily consumption of fruits was rather low. Daily consumption of citrus fruits and tropical fruits was high in India and Iran, and Iran showed higher daily consumption of pome fruits than other countries. These differences could be an example of cultural differences in eating habits among countries.

We were unable to control individual differences in the amount of each serving size of food and beverages. There are also differences in the amount of chemical ingredients even within the same plant, depending on its location and seasonal influences, and in the way the food/beverage is prepared. Another issue to take into consideration is the possibility that the results of this survey could be telling the people who consume various vegetables daily are more cautious of their overall health and may modify other activities of daily living other than their eating habits, such as self-distancing, wearing masks, and sanitizing their hands and other things. As such, there are also limits in the study using surveys.

In conclusion, our results suggest that daily consumption of phytochemical compounds included in the vegetables may have contributed to preventing contraction of COVID-19 as well as to accelerating the recover from COVID-19. The food and beverage categories that were found to be eaten daily more by the people who did not get COVID-19 are the candidates to conduct studies in detail with more controlled experimental conditions, studies using animal model systems, and at the level of phytochemical compounds to determine mechanisms of action in future.

## Data Availability

All data produced in the present study are available upon reasonable request to the authors

## Acknowledgements

We are grateful to the participants in our survey and Pierre Johane Joseph for trying the survey prior to launch.

Kang Zhu, Yanni Zhang, and Chao Yu helped Jingguo Chen in translating the survey into Chinese and spreading the survey. We would like to express our gratitude to Angela Bassoli, Michael Farruggia, Antonella Di Pizio, and Shannon Olsson for their comments to our study.

## Author contributions

SK, PVJ, VDCS, and TH served as the leadership team of the project to make decisions at each time points. All authors participated in developing the English version of the survey. The English version of the survey was translated into Chinese, Indian English, Italian, Japanese, Persian, Polish, Russian, Spanish, and Turkish, and the followings were in participated in the translation of each language: JC for Chinese, RisK and RitK for Indian English, CM-C and OC for Italian, RU and EM for Japanese, SP, RA, ZN, and FT-H for Persian, DJS for Polish, TKL, MAK, and VVV for Russian, MDG and PRD for Spanish, and EÖP and MHÖ for Turkish. All authors participated in advertising the survey. RisK, RitK, PA, HN, KWC, and SK analyzed the data. SK wrote the first draft of the paper and all authors participated in editing the draft. All authors read and approved the final manuscript.

## Availability of data and materials

## Declarations

### Ethics approval and consent to participate

The survey contained a question that asked consent to participate and we obtained consent from the participants. Ethical approval was obtained as an exempt study from Indiana University Human Research Protection Program (HRPP) in the U.S.A. (protocol #14915), the Office of Regulatory Research Compliance of Howard University (IRB-2022-0380), the Jikei University School of Medicine Ethics Committee in Japan (#34-003), the Bioethics Committee at the A.N. Severtsov Institute of Ecology & Evolution of Russian Academy of Sciences (no.2022-63-NC), and by the Second Affiliated Hospital of Xi’an Jiaotong University, Medical Ethics Committee in China (#2022023).

## Supplementary Information

### Supplementary Methods

#### Development of Survey

Questions about eating habits were constructed by asking how frequently the participants ate each category of foods, and the category was based on the food classification by the Ministry of Health Labour and Welfare of Japan (https://www.mhlw.go.jp/english/topics/foodsafety/positivelist060228/dl/r04.pdf). The food categories that we used were: citrus fruits, berries, pome fruits, stone fruits, tropical fruits, other fruits, leaf vegetables, root vegetables, other vegetables, beans and peas, nuts and seeds, herbs and spices, cereals and grains containing buckwheat and barleys, other foods such as mushrooms, and fermented foods. Buckwheat and barleys were specifically selected as representatives of cereals and grains and based on a paper showing that the phytochemicals included in them were effective on COVID-19 [15]. In order to enhance the understanding of the types of foods and the ease of answering survey questions, we listed some examples of foods in the questions pertaining to food categories. For example, in asking the frequency of eating root vegetables, we asked “Root vegetables, such as carrot, turnip, garlic, beetroots, parsnip roots, parsley root, radish, onion, sacred lotus, taro, yam, sweet potato, potato?” (English version). As the participants took the survey from around the world, it was necessary to adjust the examples depending on the language version because of the differences in the availability and familiarity of certain food items. In addition, we did not want the availability of the examples to affect the answers. There were also some food items that were named differently, depending on their location. Thus, the Indian English version was prepared separately. Questions on drinking habits were constructed by asking how frequently the participants drank four types of beverages: 1) Tea, based on the studies showing the chemical ingredients found within, had effects on coronavirus [8, 18], 2) other tea and herbal teas, not made from tea plants, 3) coffee, 4) cider, 5) hot chocolate, and 4) alcohol. Similar to the questions pertaining to foods, we showed some examples to enhance the ease of understanding. In addition, we made some adjustments to the examples shown in each language version of the survey (for example, the Russian and Turkish versions had examples of vodka and raki, respectively, instead of brandy, which other language versions may have contained). The Persian version did not have questions on cider and alcohol. The lists of these examples are shown in Supplementary Table 1.

Regarding COVID-19, we asked to choose from the choices of “Did not get COVID”, “Not get tested but think yes”, and “Tested positive”. We also asked if they got COVID-19, when they got sick and how long it took to recover (not necessarily based on PCR results as there was the “Not tested” group). We asked what symptoms they experienced as well as their severity. The symptoms that were asked included if people experienced a headache, cough, shortness of breath, runny nose or congestion, muscle/body aches/soreness, chills, lightheadedness or brain fog, dizziness, nausea, fatigue, memory loss or concussion or hallucination, diarrhea or gastrointestinal symptoms, hair loss, smell loss or reduction, taste loss or reduction, chemesthesis loss or reduction, other symptoms. Separately, we asked if they have persisting symptoms (PASC, or Long-COVID).

The English version of the questionnaire was developed first using Qualtrics software (Qualtrics, Provo, UT, USA, https://www.qualtrics.com). Subsequently, the survey was translated into nine other languages, thus getting a total of 10 language versions, namely Chinese, English, Indian English, Italian, Japanese, Persian, Polish, Russian, Spanish, and Turkish. The survey has been online since February 2022. Data used for the analysis were obtained between February 2022 and May 2023. The frequency of eating and drinking was asked following the style of the Diet History Questionnaire (DHQ) by the National Cancer Institute (NCI) [50] as a validated questionnaire for surveys on nutrition and modified the choices of answers of DHQ to “Never”, “Daily”, “Weekly”, “Monthly”. We also asked the people surveyed which type of food items and beverages they consumed using a multiple selection question. As the examples do not cover all the types of foods in each category, we also added a question where survey participants could include what food or beverage item they ate or drank.

---

**Supplementary Figure 1.**
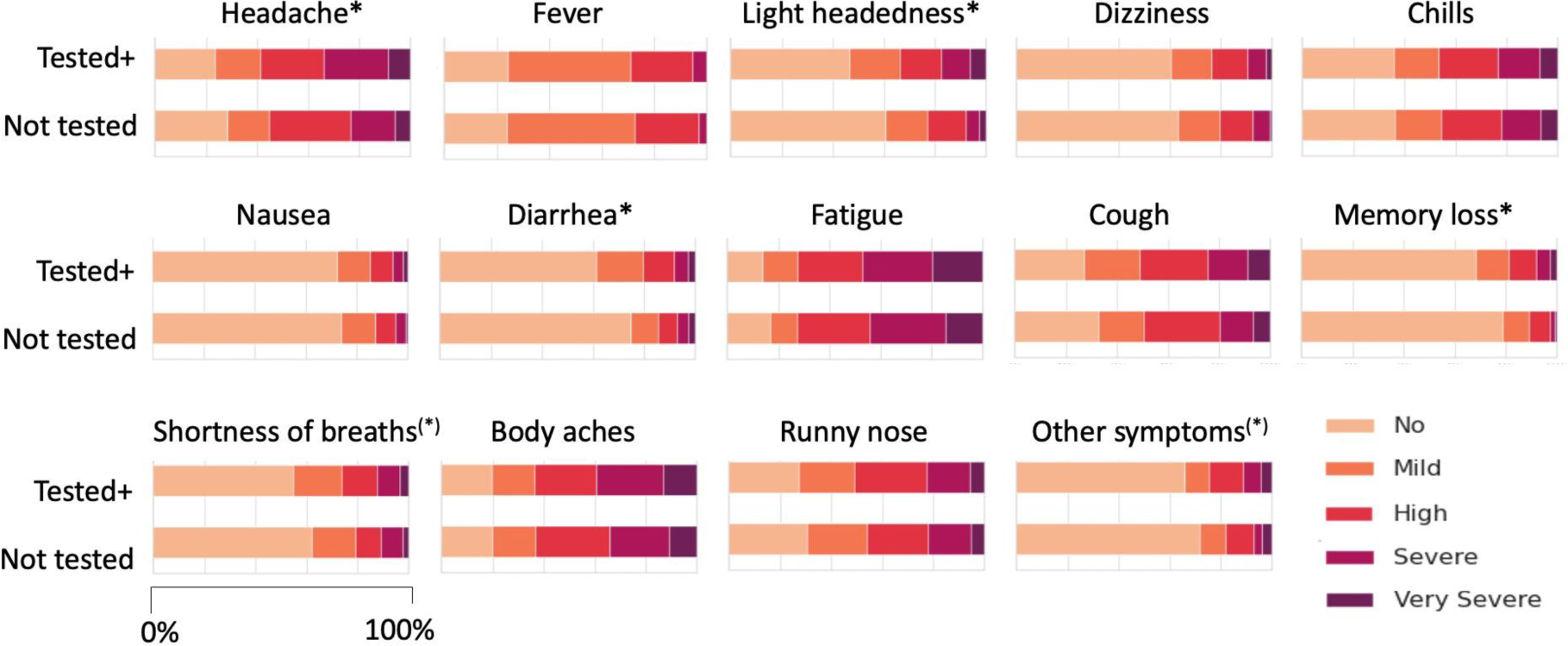
Symptoms of COVID-19 reported by the “Tested Positive” group and “Not Tested” group. *: P<0.05, (*): 0.05<P<0.10

**Supplementary Figure 2.**
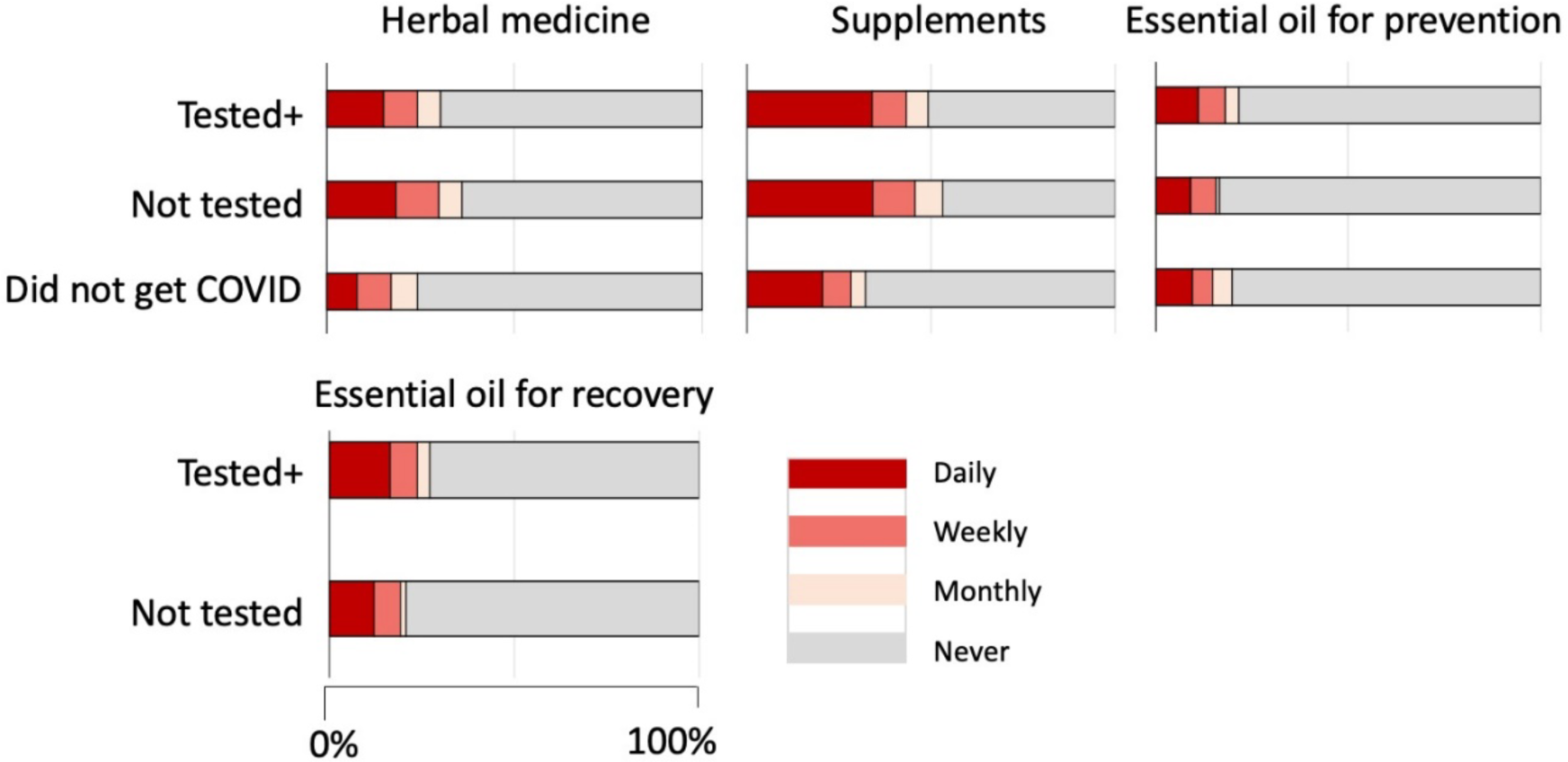
Intake of herbal medicines, supplements, and usage of essential oils.

**Supplementary Figure 3.**
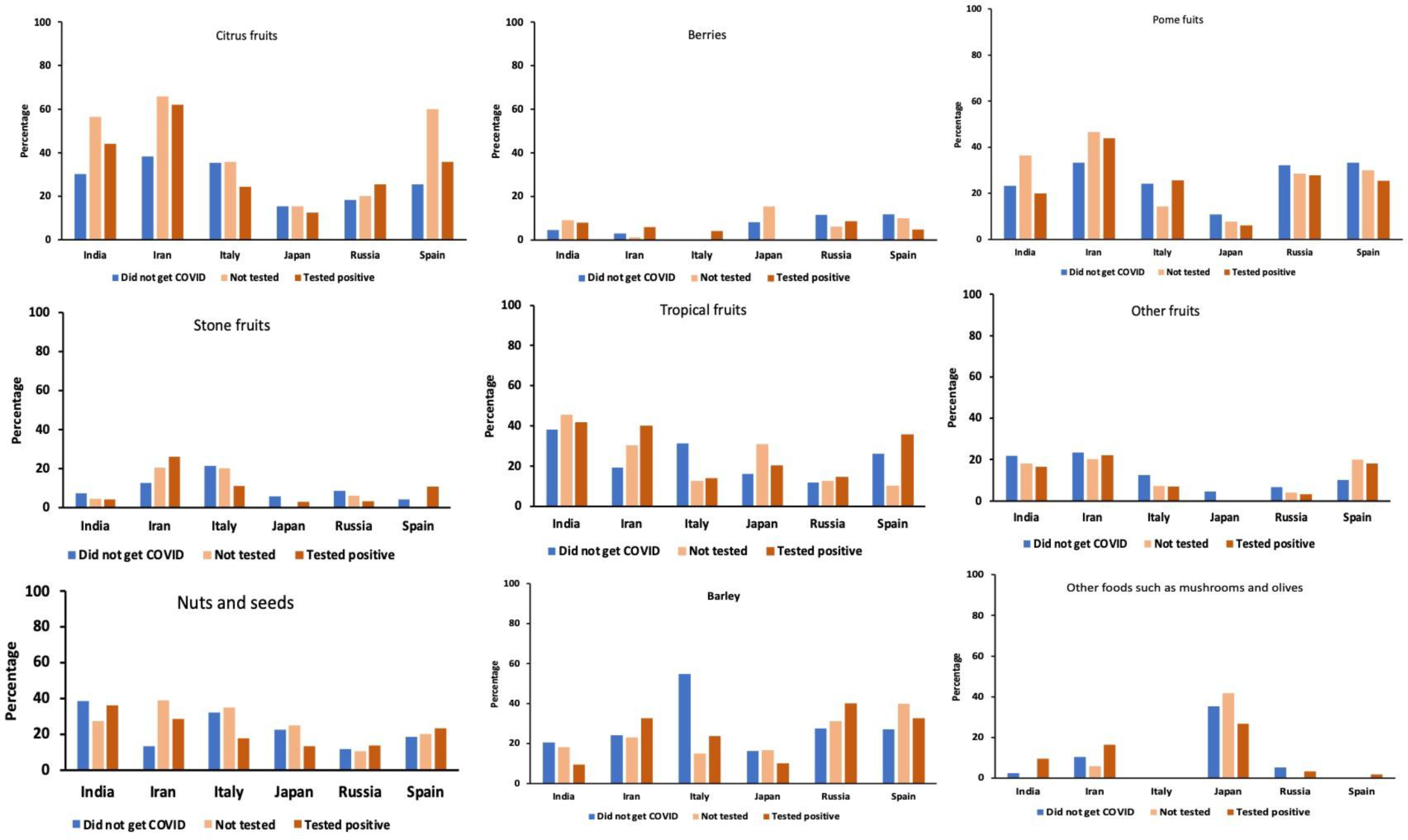
Daily intake of foods of categories other than those shown in the main text, split by countries. Blue bars: “Did not get COVID-19” group, light orange bars: “Not tested” group, Dark orange bars: “Tested positive” group

**Supplementary Figure 4.**
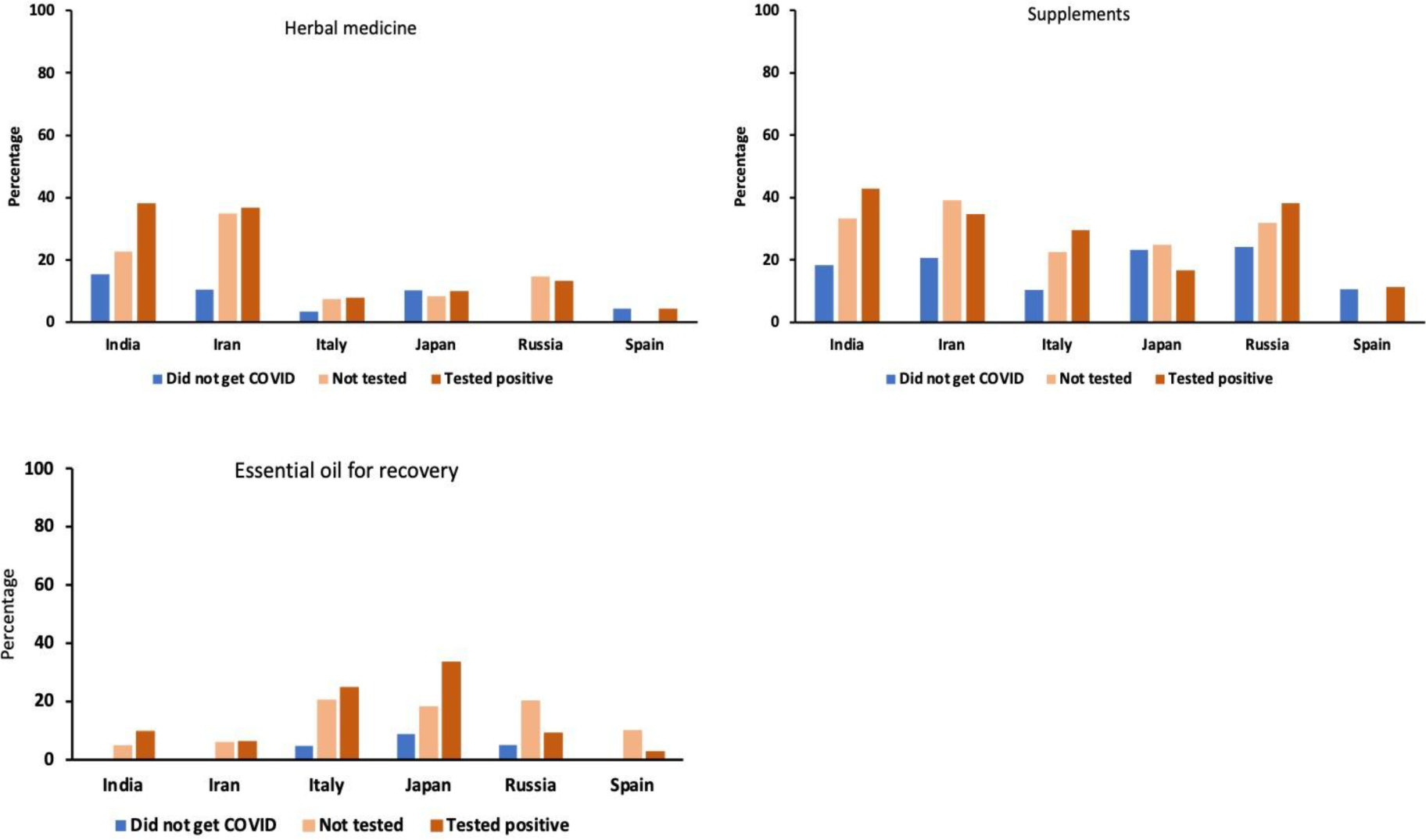
Daily intake of herbal medicine and supplements, and daily use of essential oils,split by countries.

**Supplementary Figure 5.**
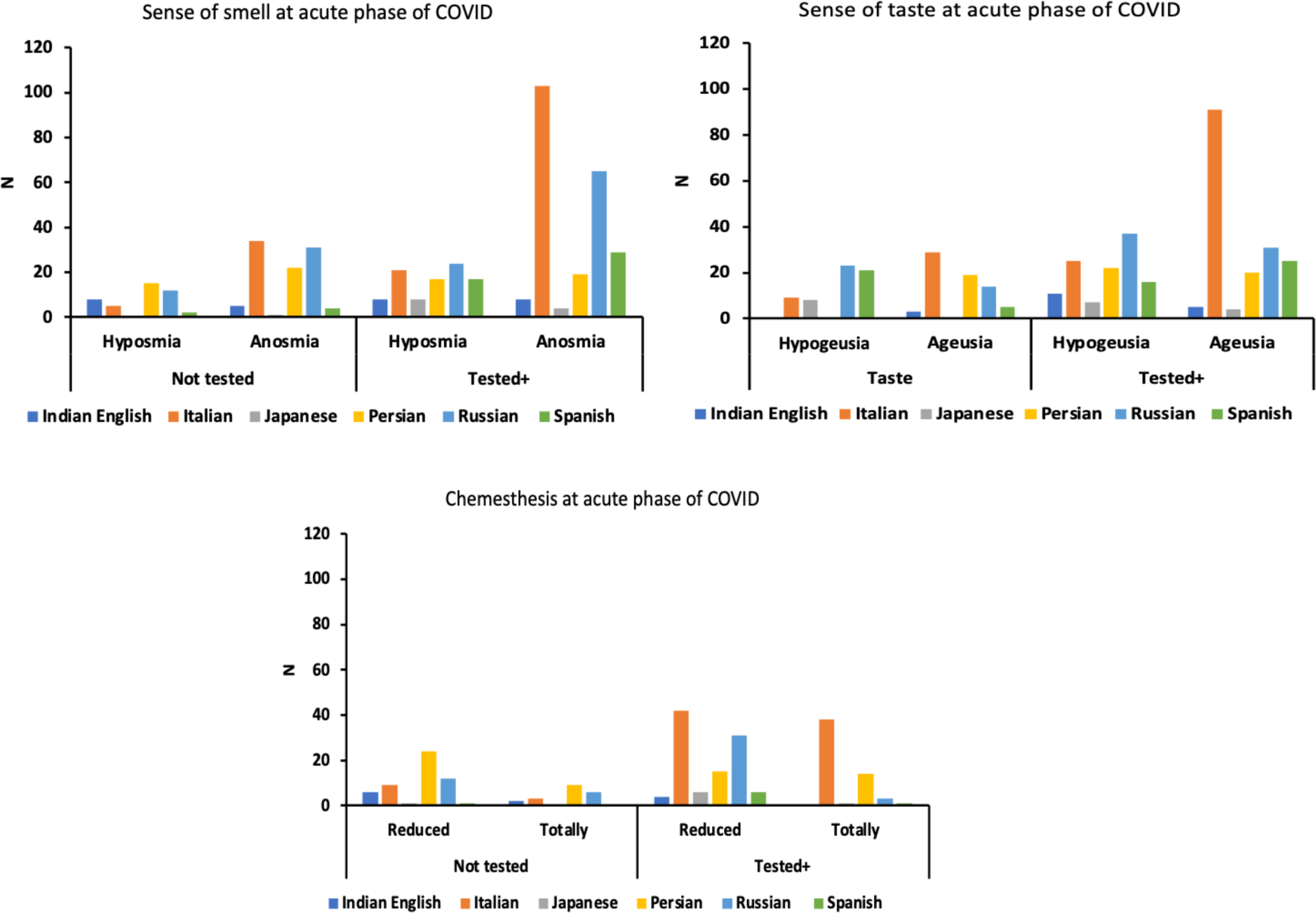
Number of participants reporting experiences of chemosensory dysfunction due to COVID-19.

**Supplementary table 1.** Examples of food items used in each language version of the survey (Part 1: Chinese, English, Indian English, and Japanese)

**Supplementary table 1.** Examples of food items used in each language version of the survey (Part 2. Persian, Polish, Russian, Spanish, and Turkish)

Supplementary Table 1 is removed from MedRxiv as it contains other languages, MedRxiv has a policy that “any information presented in a language other than English should be removed or completely replaced with the English translation”. Please contact corresponding author for access to this information.

**Supplementary Table 2.**
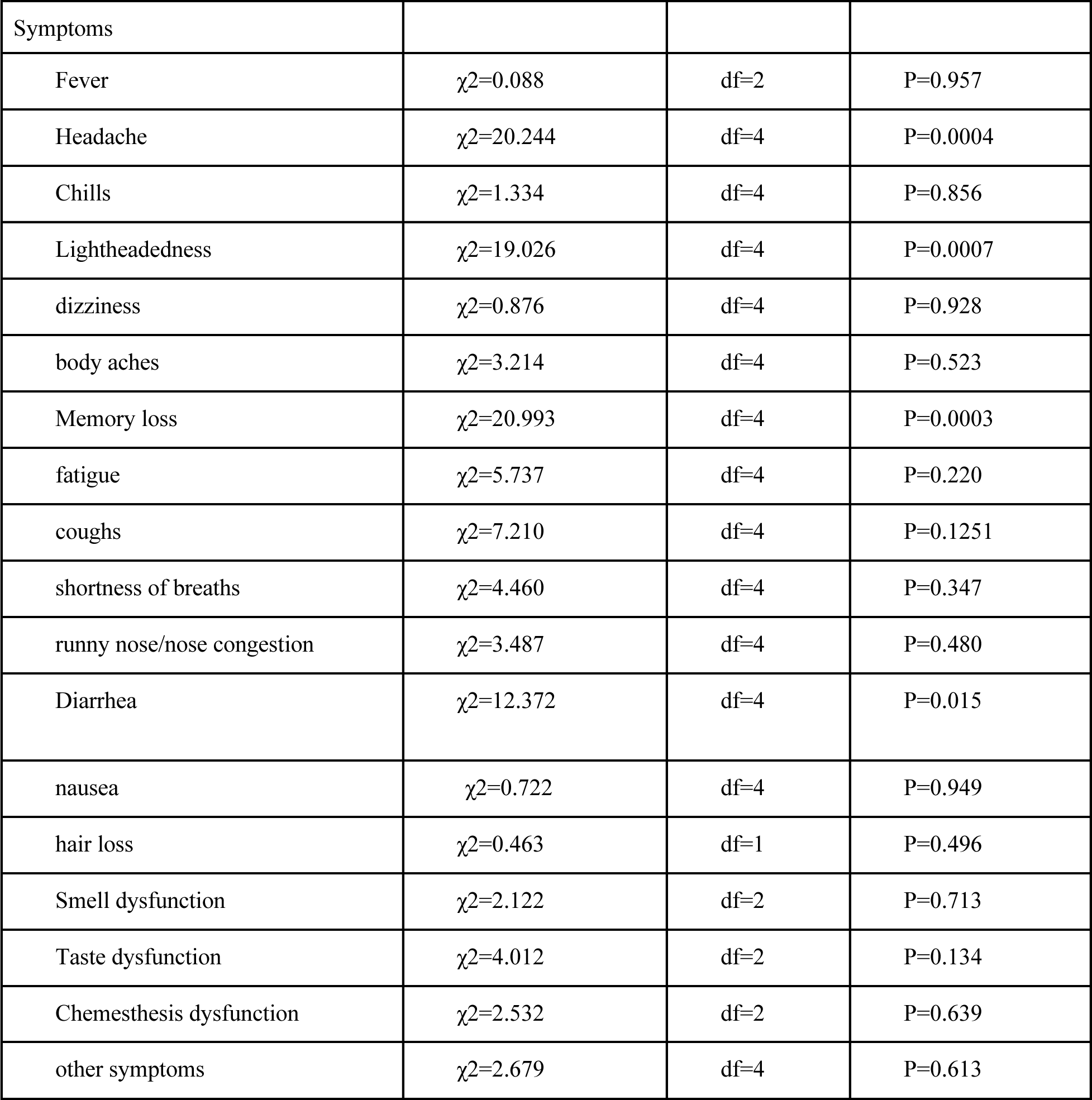
Comparison of the severity of symptoms reported by “Not tested” and “Tested positive” groups.

| Symptoms |  |  |  |
| --- | --- | --- | --- |
| Fever | $\chi^2=0.088$ | df=2 | P=0.957 |
| Headache | $\chi^2=20.244$ | df=4 | P=0.0004 |
| Chills | $\chi^2=1.334$ | df=4 | P=0.856 |
| Lightheadedness | $\chi^2=19.026$ | df=4 | P=0.0007 |
| dizziness | $\chi^2=0.876$ | df=4 | P=0.928 |
| body aches | $\chi^2=3.214$ | df=4 | P=0.523 |
| Memory loss | $\chi^2=20.993$ | df=4 | P=0.0003 |
| fatigue | $\chi^2=5.737$ | df=4 | P=0.220 |
| coughs | $\chi^2=7.210$ | df=4 | P=0.1251 |
| shortness of breaths | $\chi^2=4.460$ | df=4 | P=0.347 |
| runny nose/nose congestion | $\chi^2=3.487$ | df=4 | P=0.480 |
| Diarrhea | $\chi^2=12.372$ | df=4 | P=0.015 |
| nausea | $\chi^2=0.722$ | df=4 | P=0.949 |
| hair loss | $\chi^2=0.463$ | df=1 | P=0.496 |
| Smell dysfunction | $\chi^2=2.122$ | df=2 | P=0.713 |
| Taste dysfunction | $\chi^2=4.012$ | df=2 | P=0.134 |
| Chemesthesis dysfunction | $\chi^2=2.532$ | df=2 | P=0.639 |
| other symptoms | $\chi^2=2.679$ | df=4 | P=0.613 |

**Supplementary Table 3.**
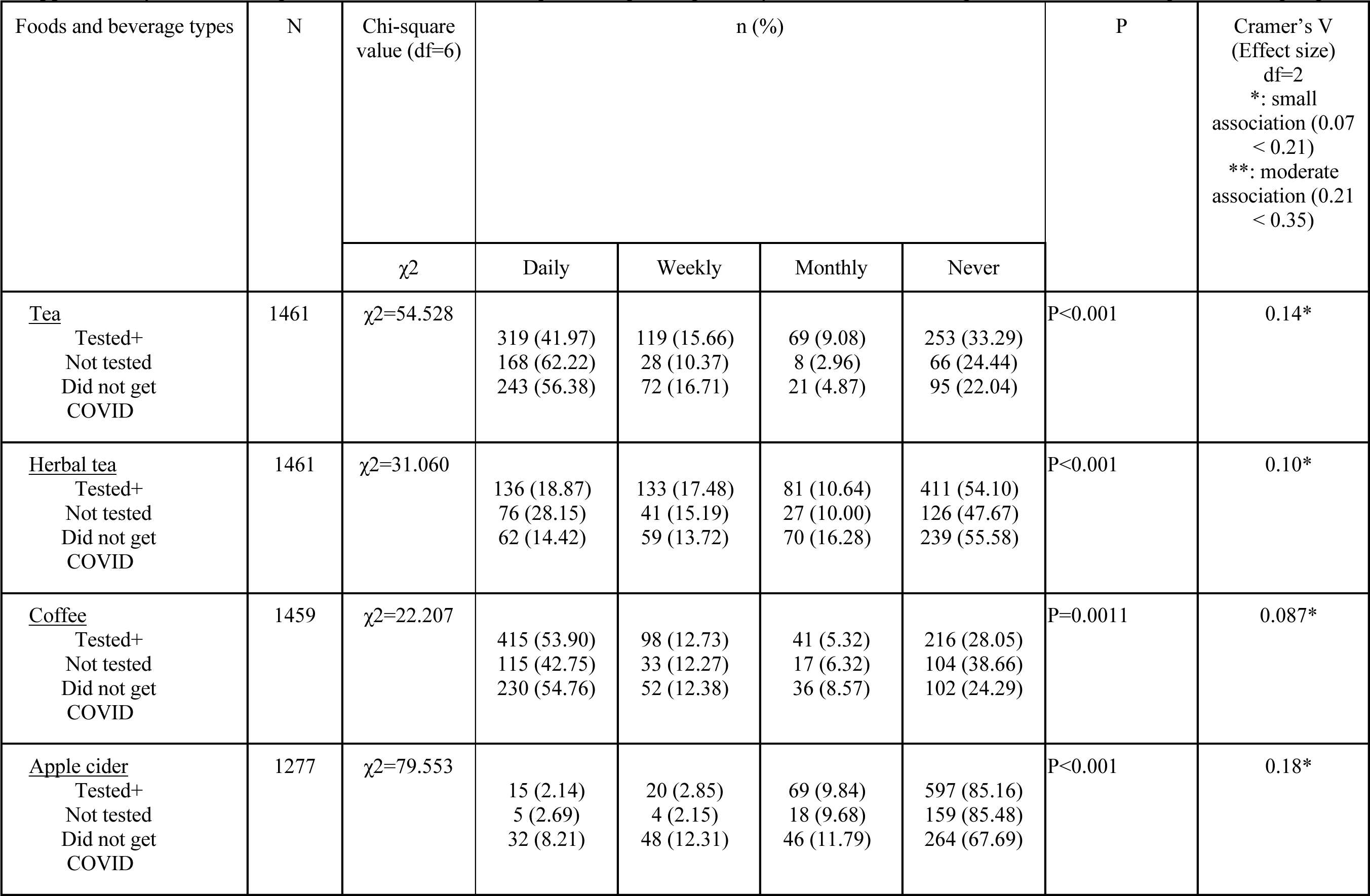
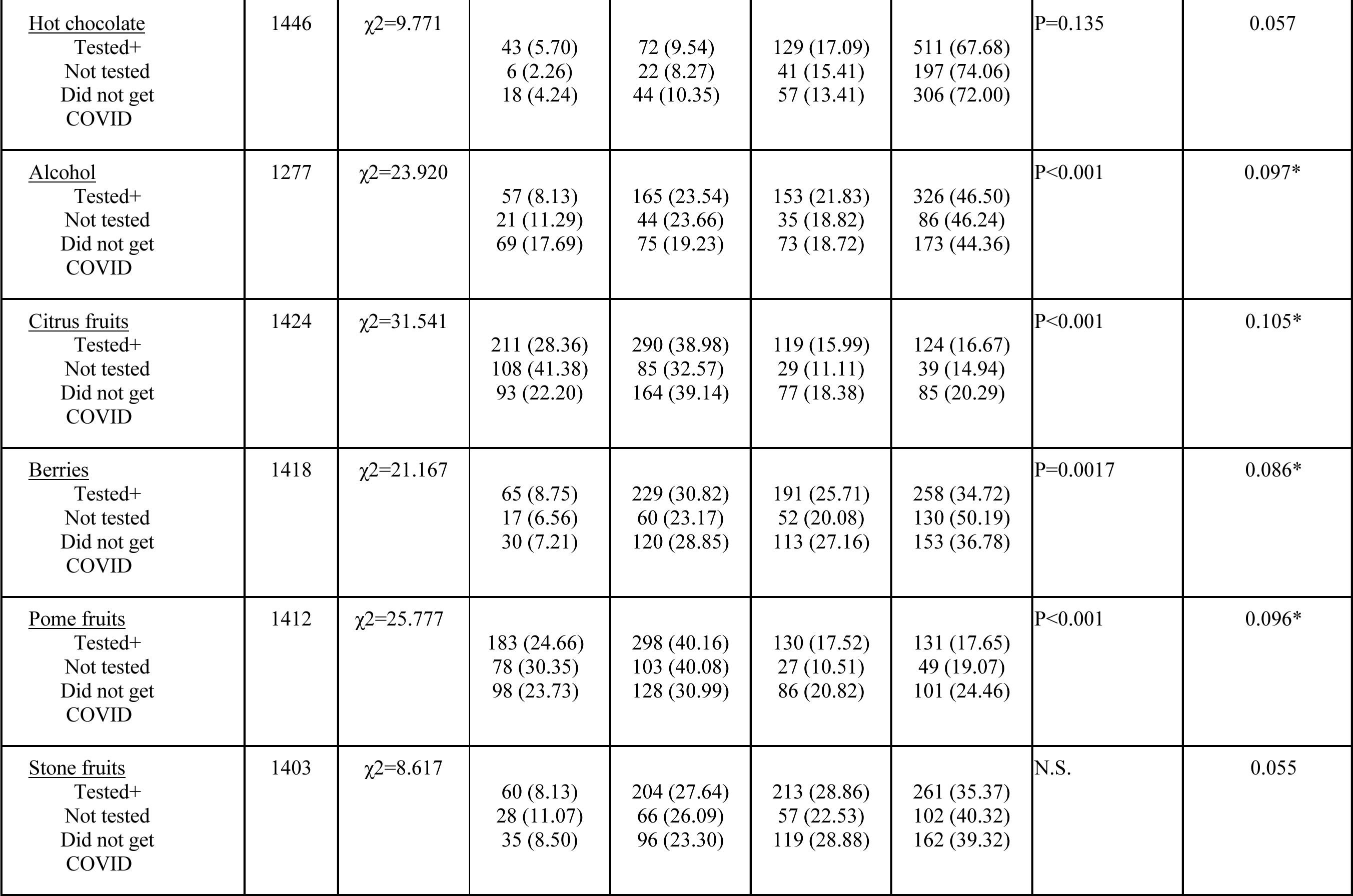
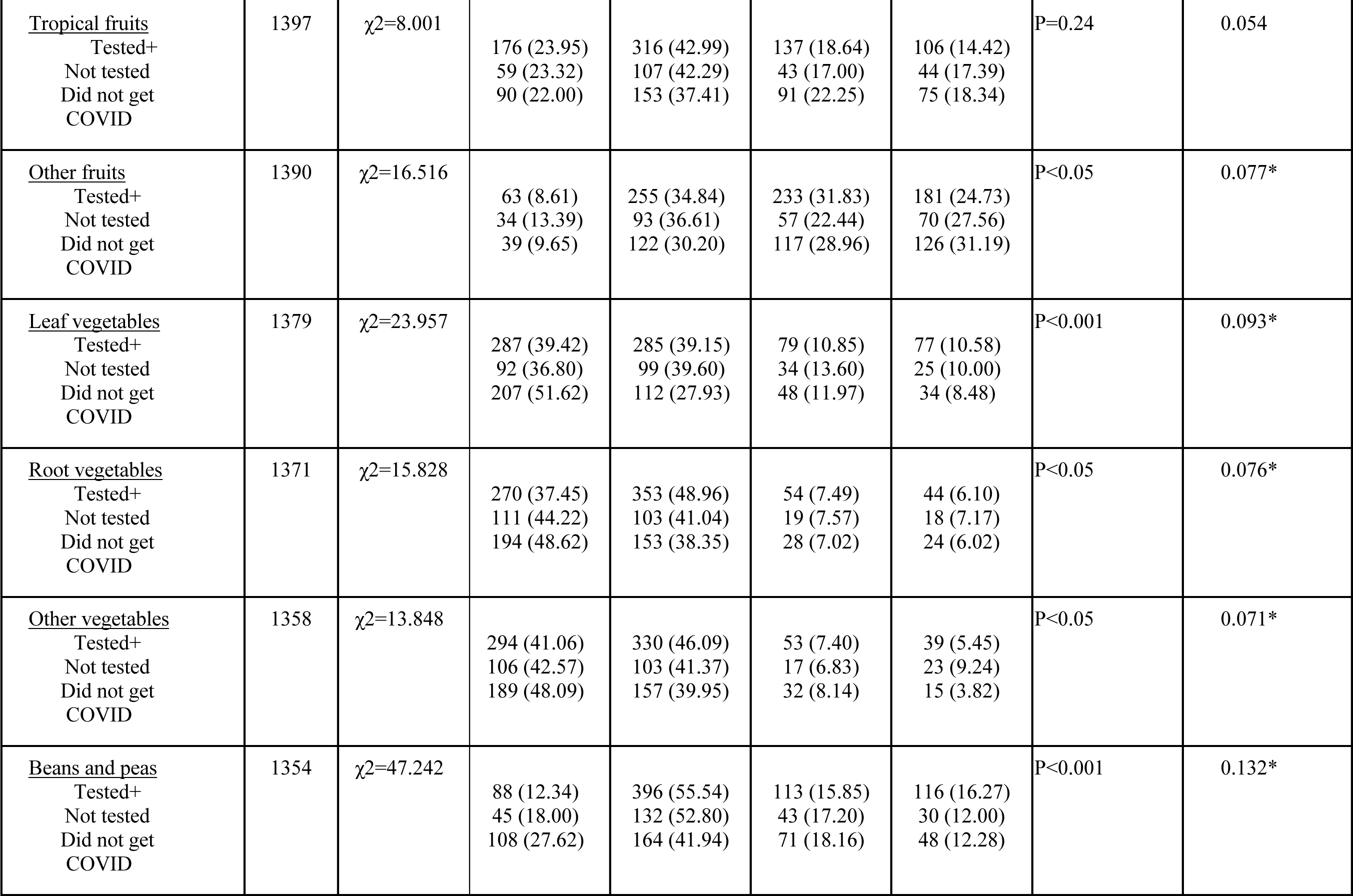
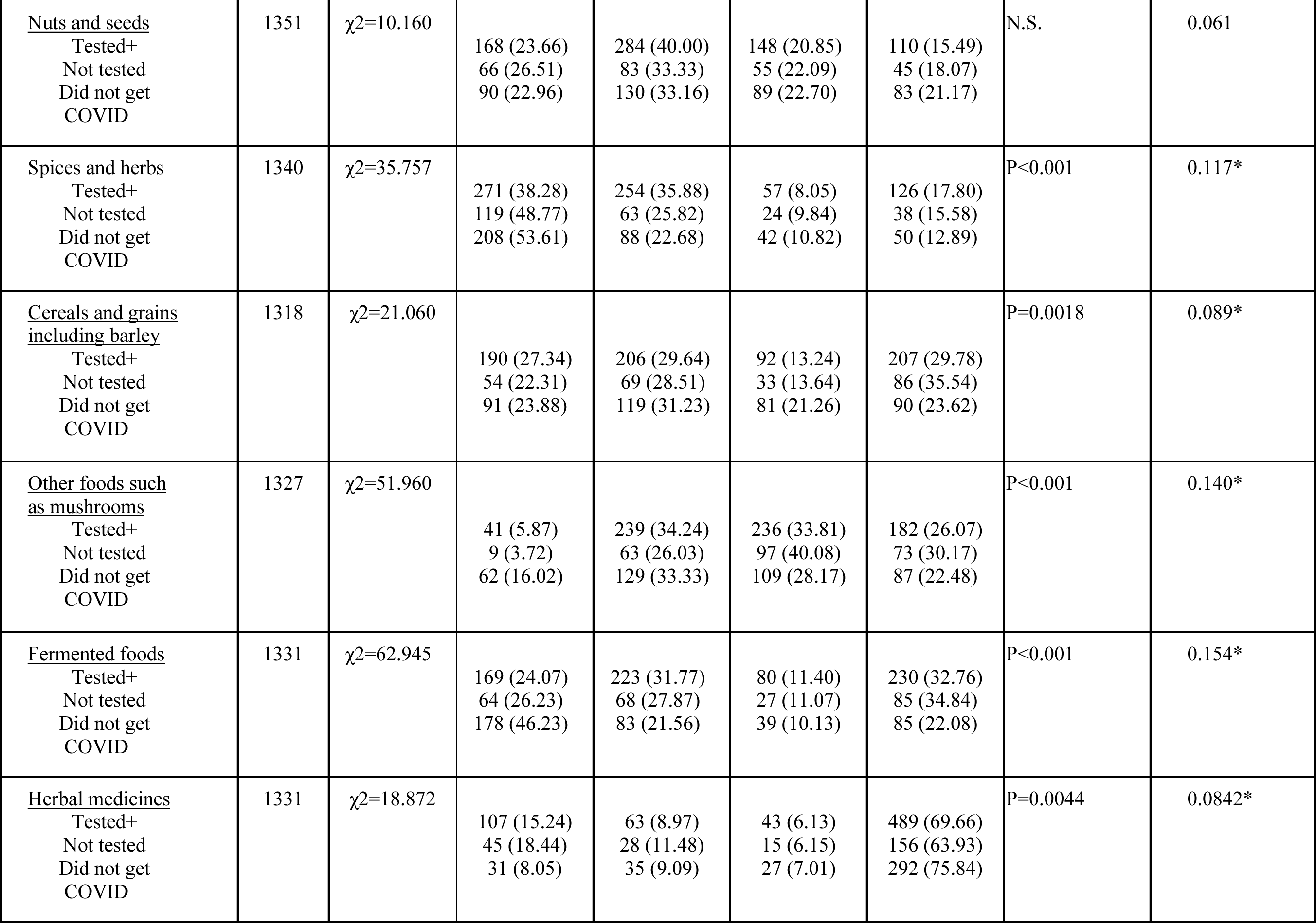
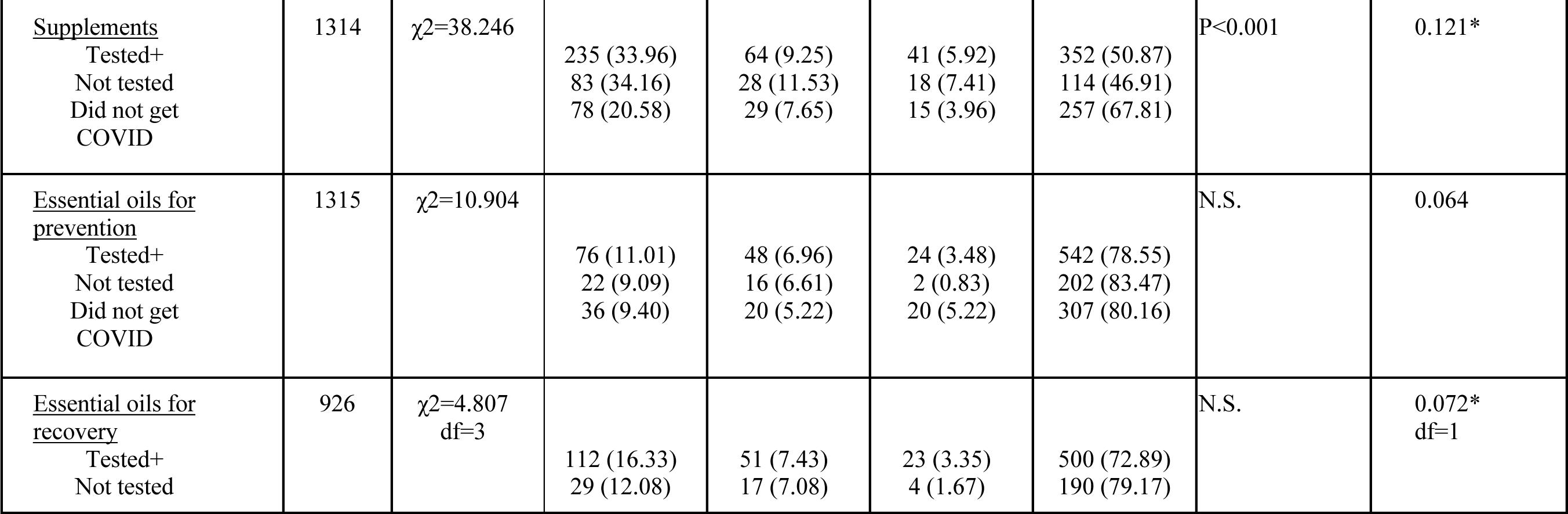
Comparison of the food and beverage consumption reported by “Not tested”, “Tested positive”, and “Did not get COVID” groups.

**Supplementary Table 4.**
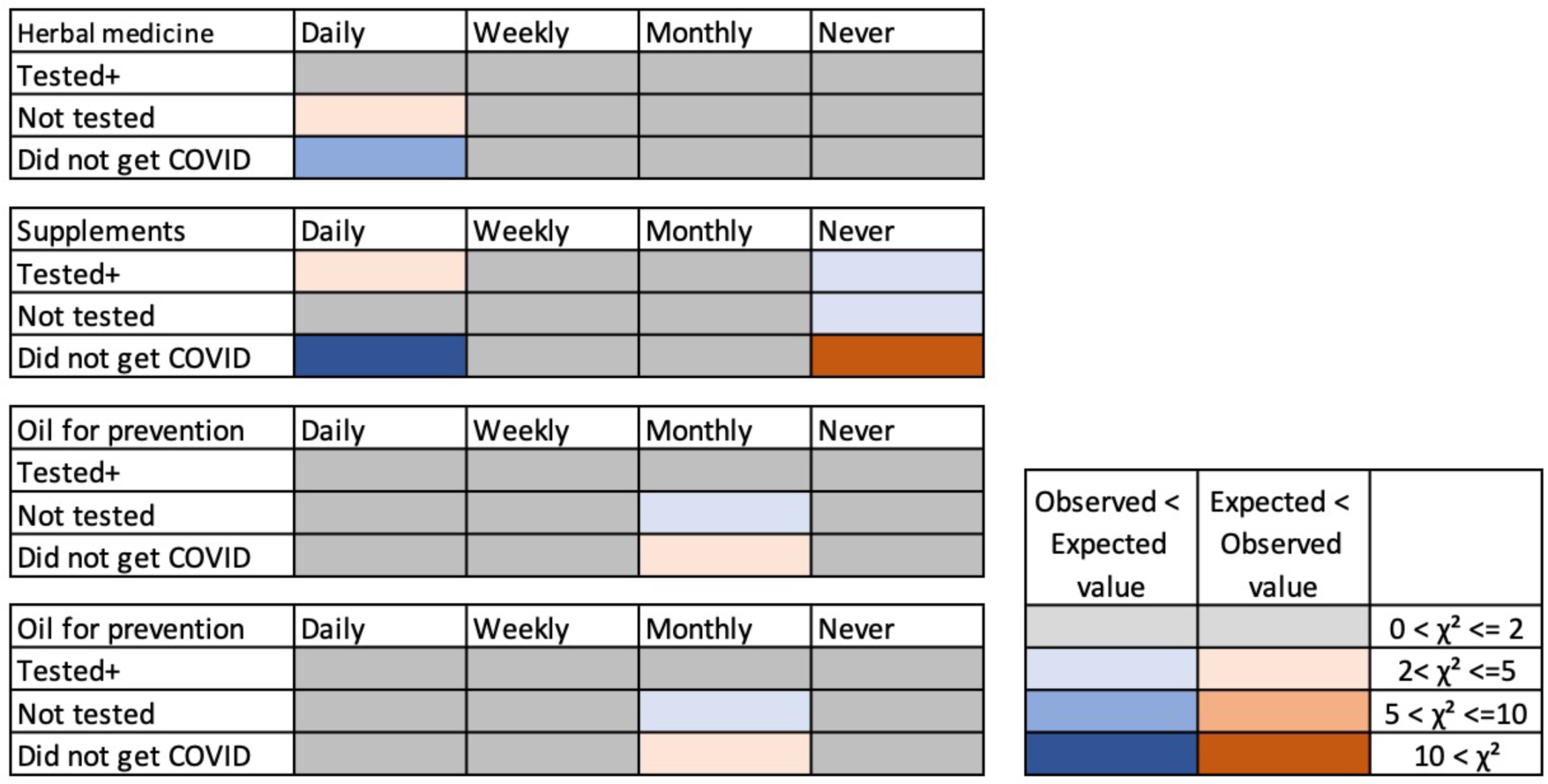
Discrepancy from expected values. The colors indicate where the observed values were higher than the expected values (smaller to larger discrepancy indicated by pink to red color), and the observed values were lower than the expected values (smaller to larger discrepancy indicated by light to dark blue). Grey color indicates there were no or negligible discrepancies: Results on herbal medicines, supplements, essential oils.

**Supplementary Table 5.**
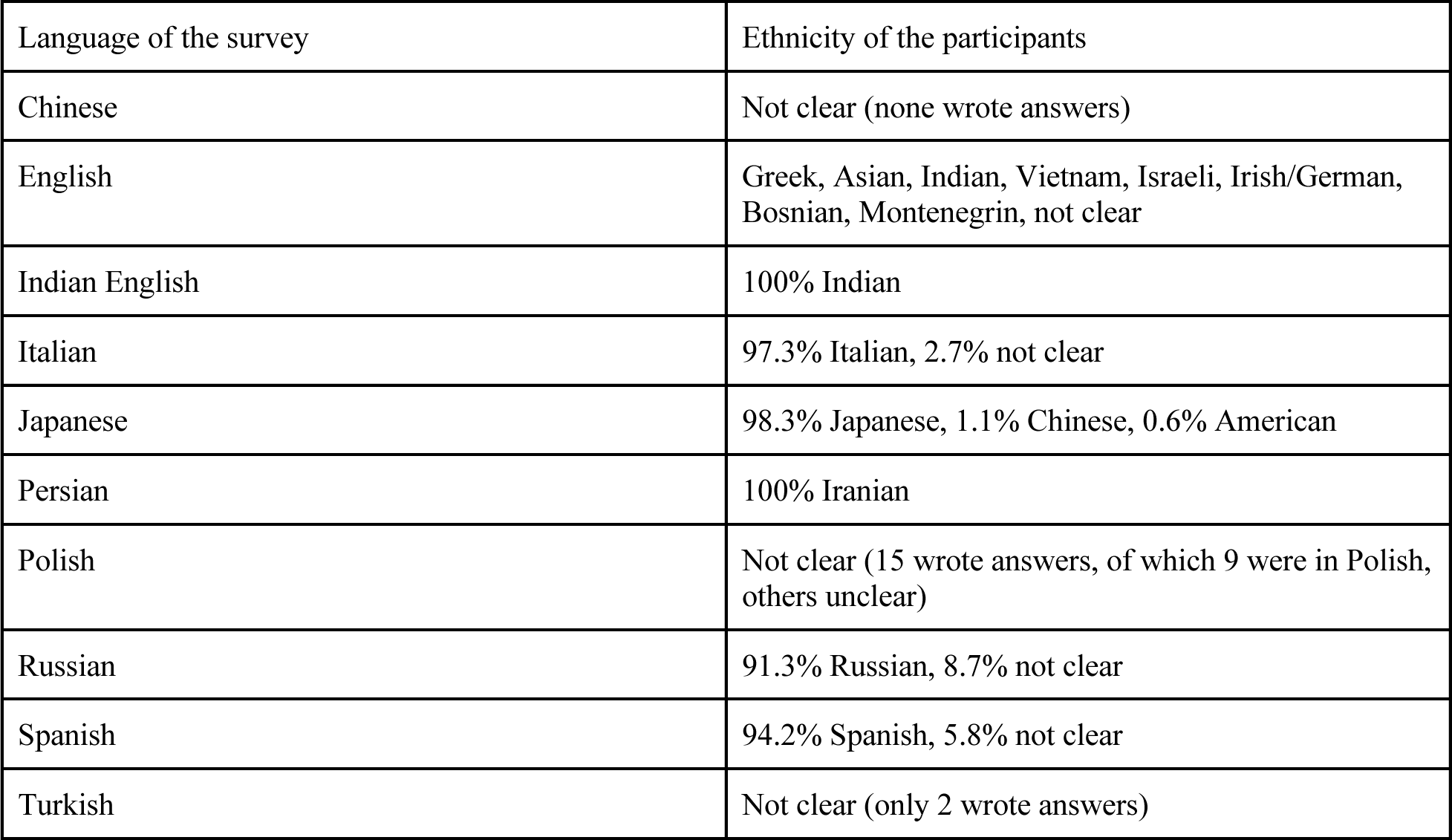
Ethnicity of the participants who answered each language version of the survey.

## Supplementary Material

### English version of the survey

GCCR - Phytochemicals - English1

Q82 I am at least 18 years old, and I wish to voluntarily participate in this study.

- Yes (1)
- No (2)

Skip To: End of Survey If I am at least 18 years old, and I wish to voluntarily participate in this study. = No End of Block: Block 1

Start of Block: Block 2

During the pandemic, many people have tried various methods to help accelerate their recovery from symptoms of COVID-19. Among the most popular approaches are at home remedies and herbal medicines. It is still unclear whether some foods, drinks, essential oils, and their ingredients may be helpful in recovering from COVID-19.

In this survey, we would like to hear if you tried any specific foods, drinks, home remedies, or herbal medicines that you have heard or think work in suppressing COVID-19 symptoms and/or in facilitating recovery. If you did, please let us know – we would like to hear their effects.

Thank you so much for your voluntary participation. The data will be de-identified, saved and protected in our system.

– Global Consortium for Chemosensory Research Team

Please let us know if you have any questions regarding the study.

General contact: Dr. Sachiko Koyama, Indiana University, +1-812-345-6155

One of the lead researchers has a company that does research and development of products using phytochemicals. This company is not funding or sponsoring this research study. We are giving you this information so you can decide if this affects your willingness to participate in this study.

Q2 Did you get COVID-19?

- Not tested, but I suspect that I got it (1)
- Yes – tested positive via PCR or antibody test (2)
- No or not aware (3)

Display This Question:

If Did you get COVID-19? = Not tested, but I suspect that I got it

Or Did you get COVID-19? = Yes – tested positive via PCR or antibody test

Q3 If yes, what symptoms did/do you have (overall and when at worst)?

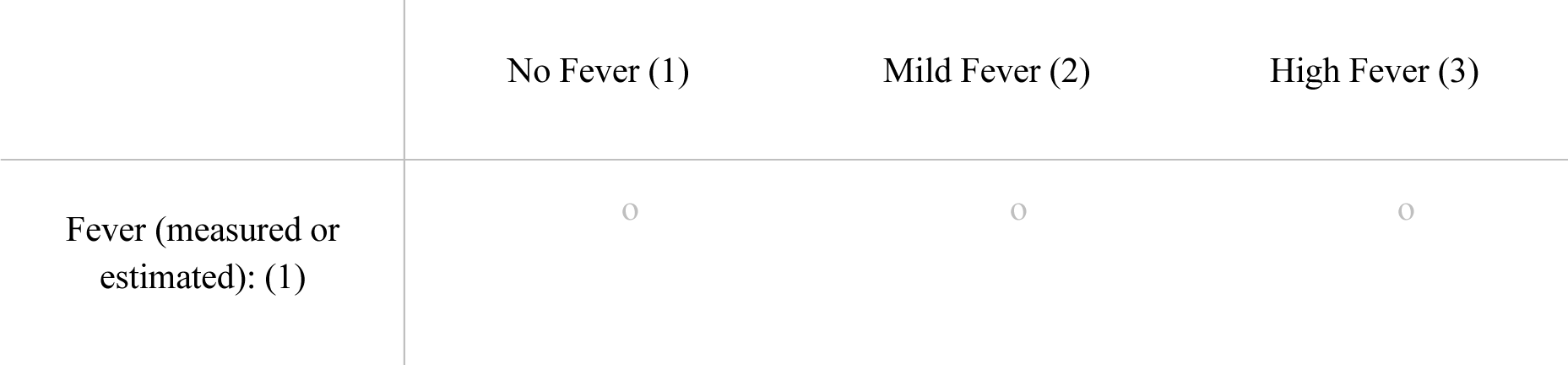

Display This Question:

If Did you get COVID-19? = Not tested, but I suspect that I got it

Or Did you get COVID-19? = Yes – tested positive via PCR or antibody test

Q4 (Continued)

|  | No (1) | Mild (2) | Moderate (3) | Severe (4) | Very Severe (5) |
| --- | --- | --- | --- | --- | --- |
| Headache (1) | ☐ | ☐ | ☐ | ☐ | ☐ |
| Cough (2) | ☐ | ☐ | ☐ | ☐ | ☐ |
| Shortness of<br>breath (3) | ☐ | ☐ | ☐ | ☐ | ☐ |
| Runny nose or<br>congestion (4) | ☐ | ☐ | ☐ | ☐ | ☐ |
| Muscle/body<br>aches/soreness (5) | ☐ | ☐ | ☐ | ☐ | ☐ |
| Chills (6) | ☐ | ☐ | ☐ | ☐ | ☐ |
| Light headedness<br>or brain fog (7) | ☐ | ☐ | ☐ | ☐ | ☐ |
| Dizziness (8) | ☐ | ☐ | ☐ | ☐ | ☐ |
| Nausea (9) | ☐ | ☐ | ☐ | ☐ | ☐ |
| Fatigue (10) | ☐ | ☐ | ☐ | ☐ | ☐ |
| Memory loss,<br>confusion,<br>hallucination (11) | ☐ | ☐ | ☐ | ☐ | ☐ |
| Diarrhea,<br>gastrointestinal<br>symptoms (12) | ☐ | ☐ | ☐ | ☐ | ☐ |
| Other symptoms<br>(13) | ☐ | ☐ | ☐ | ☐ | ☐ |

Display This Question:

If Did you get COVID-19? = Not tested, but I suspect that I got it

Or Did you get COVID-19? = Yes – tested positive via PCR or antibody test

Q5 (Continued)

|  | Yes (1) | No (2) |
| --- | --- | --- |
| Hair loss (1) | ☐ | ☐ |

Display This Question:

If Did you get COVID-19? = Not tested, but I suspect that I got it

Or Did you get COVID-19? = Yes – tested positive via PCR or antibody test

Q6 (Continued)

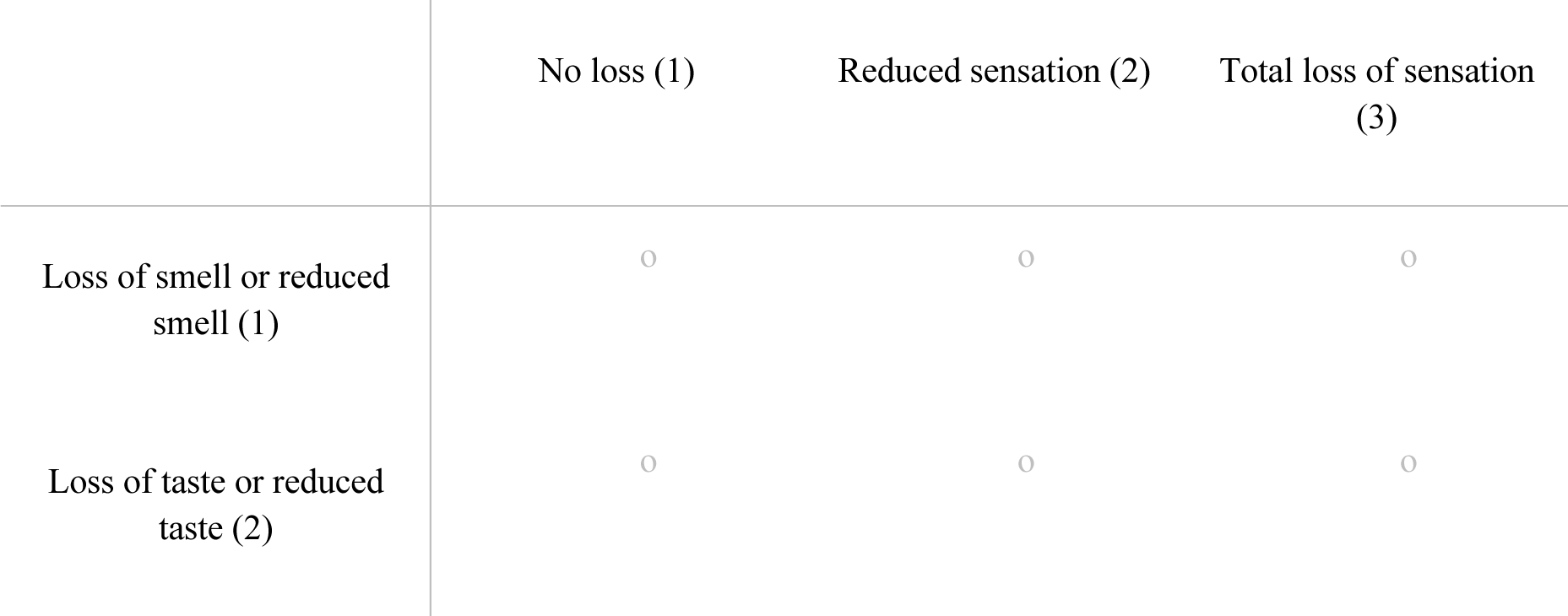
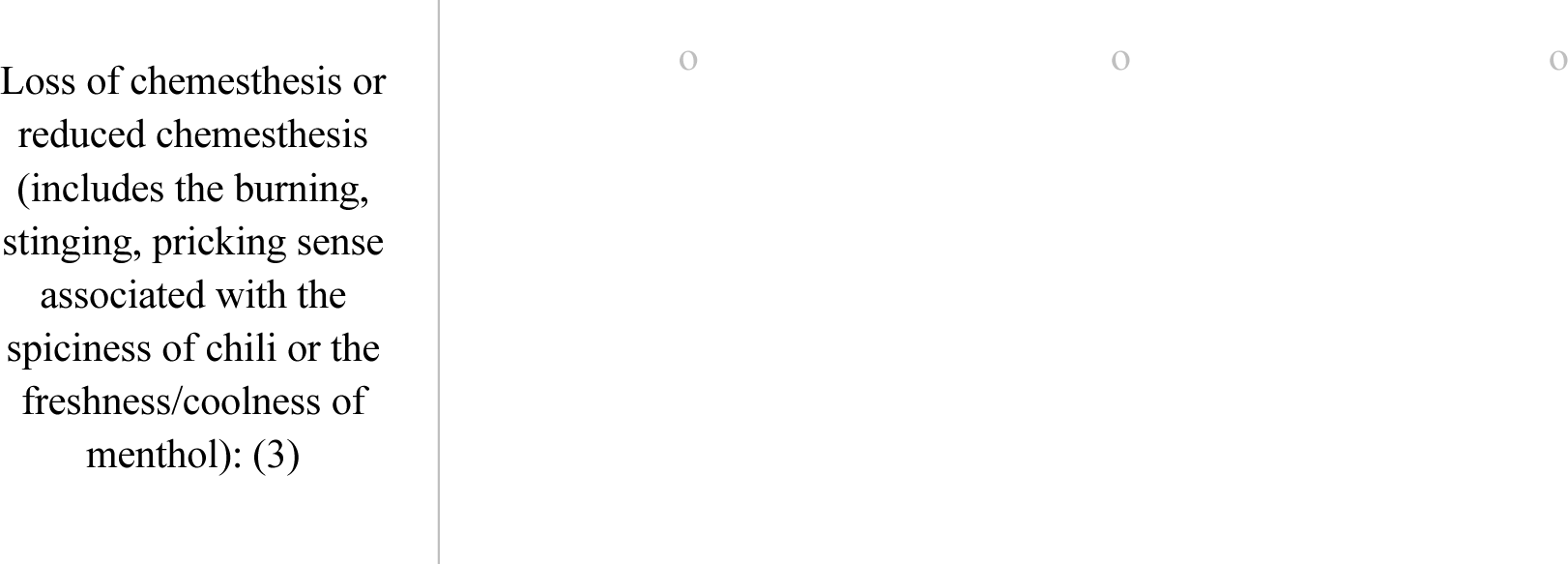

Display This Question:

If (Continued) = Loss of taste or reduced taste [Reduced sensation]

Or (Continued) = Loss of taste or reduced taste [Total loss of sensation]

Q7 If you experienced reduced ability to taste or total loss of taste, which sensation?

▢ Sweet (1)
▢ Sour (2)
▢ Salty (3)
▢ Bitter (4)
▢ Umami (savory) (5)

Display This Question:

If Did you get COVID-19? = Yes – tested positive via PCR or antibody test

Or Did you get COVID-19? = Not tested, but I suspect that I got it

Q8 When did you have COVID for the first time (in case you got it multiple times)?

- Before February 2020 (1)
- Between March to May, 2020 (2)
- Between June to August, 2020 (3)
- Between September to November, 2020 (4)
- Between December 2020 to February 2021 (5)
- Between March to May, 2021 (6)
- June to August, 2021 (7)
- September to November, 2021 (8)
- After December 2021 (9)

Display This Question:

If Did you get COVID-19? = Yes – tested positive via PCR or antibody test

Or Did you get COVID-19? = Not tested, but I suspect that I got it

Q9 Approximately how many days did it take to have a negative COVID test, or how many days did it take to recover from the worst phase (if you did not get tested)?

Display This Question:

If Did you get COVID-19? = Yes – tested positive via PCR or antibody test

Or Did you get COVID-19? = Not tested, but I suspect that I got it

Q10 Do (did) you have persisting symptoms of COVID-19?

- Yes (1)
- No (2)

Display This Question:

If Do (did) you have persisting symptoms of COVID-19? = Yes

Q12 Which persisting symptoms do you have? Choose all that apply

▢ Fever (1)
▢ Headache (2)
▢ Cough (3)
▢ Shortness of breath (4)
▢ Runny nose or congestion (5)
▢ Muscle/body aches/soreness (6)
▢ Chills (7)
▢ Light headedness or brain fog (8)
▢ Dizziness (9)
▢ Nausea (10)
▢ Fatigue (11)
▢ Hair loss (12)
▢ Memory loss (13)
▢ Confusion (14)
▢ Hallucination (15)
▢ Loss or reduced smell (16)
▢ Loss or reduced taste (17)
▢ Loss or reduced chemesthesis (18)
▢ Diarrhea (19)
▢ Gastrointestinal Symptoms (20)
▢ Others (21)

Q13 Please answer to the following questions about your fluids/beverage consumption during the pandemic (since December 2019).

Q14 During the pandemic, how often did you drink tea such as black tea, green tea, white tea, yellow tea, oolong tea, red tea, others? (Note: This excludes Kombucha. Please check one answer that best describes the frequency of your use of the food category)

- Never (1)
- Daily (2)
- Weekly (3)
- Monthly (4)

Q15 During the pandemic, how often did you drink herbal tea such as ginger, lemongrass, licorice, others?

- Never (1)
- Daily (2)
- Weekly (3)
- Monthly (4)

Display This Question:

If During the pandemic, how often did you drink herbal tea such as ginger, lemongrass, licorice, oth… = Daily

Or During the pandemic, how often did you drink herbal tea such as ginger, lemongrass, licorice, oth… = Weekly

Or During the pandemic, how often did you drink herbal tea such as ginger, lemongrass, licorice, oth… = Monthly

Q16 What type of herbal tea do/did you drink?

▢ Ginger (11)
▢ Lemongrass (14)
▢ Others (15)

Q17 During the pandemic, how often did you drink coffee?

- Never (1)
- Daily (2)
- Weekly (3)
- Monthly (4)

Display This Question:

If During the pandemic, how often did you drink coffee? = Daily

Or During the pandemic, how often did you drink coffee? = Weekly

Or During the pandemic, how often did you drink coffee? = Monthly

Q18 What type of coffee do/did you drink?

Q19 During the pandemic, how often did you drink cocoa?

- Never (1)
- Daily (2)
- Weekly (3)
- Monthly (4)

Q20 During the pandemic, how often did you drink cider?

- Never (1)
- Daily (2)
- Weekly (3)
- Monthly (4)

Q21 During the pandemic, how often did you drink alcohol such as wine, beer, whiskey, brandy, others?

- Never (1)
- Daily (2)
- Weekly (3)
- Monthly (4)

Display This Question:

If During the pandemic, how often did you drink alcohol such as wine, beer, whiskey, brandy, others? = Daily

Or During the pandemic, how often did you drink alcohol such as wine, beer, whiskey, brandy, others? = Weekly

Or During the pandemic, how often did you drink alcohol such as wine, beer, whiskey, brandy, others? = Monthly

Q22 What type of alcoholic beverage do/did you drink? You may write more than one.

▢ beer (4)
▢ brandy (5)
▢ whiskey (6)
▢ wine (7)
▢ Others (8)

Q77 Did you change your fluid/beverage habits after the pandemic started?

- Yes (1)
- No (2)

Display This Question:

If Did you change your fluid/beverage habits after the pandemic started? = Yes

Q80 Why did you change your fluid/beverage habits?

▢ To prevent contracting the virus, I started to drink medicinal/herbal tea more often. (1)
▢ Regular liquids did not taste good anymore. (2)
▢ No specific reason (3)
▢ Other reason (4)

Display This Question:

If Did you change your fluid/beverage habits after the pandemic started? = Yes

Q78 Name of the drink

Q23 Please tell us if there are specific foods that you used during this pandemic. Fruits (check one answer that best describes the frequency of your use of the food category)

Q24 Citrus fruits such as mandarin orange, clementine, tangerine, lemon, lime, pummelo, kumquat, sweet orange?

- Never (1)
- Daily (2)
- Weekly (3)
- Monthly (4)

Display This Question:

If Citrus fruits such as mandarin orange, clementine, tangerine, lemon, lime, pummelo, kumquat, swee… = Daily

Or Citrus fruits such as mandarin orange, clementine, tangerine, lemon, lime, pummelo, kumquat, swee… = Weekly

Or Citrus fruits such as mandarin orange, clementine, tangerine, lemon, lime, pummelo, kumquat, swee… = Monthly

Q25 What citrus fruit do/did you eat?

▢ clementine (4)
▢ kumquat (5)
▢ lemon (6)
▢ lime (7)
▢ mandarin orange (8)
▢ pummelo (9)
▢ sweet orange (10)
▢ tangerine (11)
▢ others (12)

Q26 Berries such as cranberry, blueberry, huckleberry, strawberry, blackberry, raspberry, black elderberry, gooseberry, black mulberry, pitanga, black currants (cassis)?

- Never (1)
- Daily (2)
- Weekly (3)
- Monthly (4)

Display This Question:

If Berries such as cranberry, blueberry, huckleberry, strawberry, blackberry, raspberry, black elder… = Daily

Or Berries such as cranberry, blueberry, huckleberry, strawberry, blackberry, raspberry, black elder… = Weekly

Or Berries such as cranberry, blueberry, huckleberry, strawberry, blackberry, raspberry, black elder… = Monthly

Q27 What berries do you eat?

▢ blackberry (4)
▢ black currants (cassis) (5)
▢ black elderberry (6)
▢ black mulberry (7)
▢ blueberry (8)
▢ cranberry (9)
▢ gooseberry (10)
▢ huckleberry (11)
▢ pitanga (12)
▢ raspberry (13)
▢ strawberry (14)
▢ others (15)

Q84 Pome fruits such as apple, pear, quince?

- Never (1)
- Daily (2)
- Weekly (3)
- Monthly (4)

Display This Question:

If Pome fruits such as apple, pear, quince? = Daily

Or Pome fruits such as apple, pear, quince? = Weekly

Or Pome fruits such as apple, pear, quince? = Monthly

Q85 What pome fruits did you eat?

▢ apple (4)
▢ pear (5)
▢ quince (6)
▢ others (7)

Q86 Stone fruits such as plum, cherry, apricot, peach?

- Never (1)
- Daily (2)
- Weekly (3)
- Monthly (4)

Display This Question:

If Stone fruits such as plum, cherry, apricot, peach? = Daily

Or Stone fruits such as plum, cherry, apricot, peach? = Weekly

Or Stone fruits such as plum, cherry, apricot, peach? = Monthly

Q87 What stone fruits did you eat?

▢ apricot (4)
▢ cherry (5)
▢ peach (6)
▢ plum (7)
▢ others (8)

Q28 Tropical fruits such as banana, mango, papaya, kiwi, pineapple, date, avocado, soursop?

- Never (1)
- Daily (2)
- Weekly (3)
- Monthly (4)

Display This Question:

If Tropical fruits such as banana, mango, papaya, kiwi, pineapple, date, avocado, soursop? = Daily

Or Tropical fruits such as banana, mango, papaya, kiwi, pineapple, date, avocado, soursop? = Weekly

Or Tropical fruits such as banana, mango, papaya, kiwi, pineapple, date, avocado, soursop? = Monthly

Q29 What tropical fruit do/did you eat? ()

▢ avocado (4)
▢ banana (5)
▢ date (6)
▢ kiwi (7)
▢ mango (8)
▢ papaya (9)
▢ pineapple (10)
▢ soursop (11)
▢ others (12)

Q30 Other fruits such as fig, muskmelon, watermelon, and grapes?

- Never (1)
- Daily (2)
- Weekly (3)
- Monthly (4)

Display This Question:

If Other fruits such as fig, muskmelon, watermelon, and grapes? = Daily

Or Other fruits such as fig, muskmelon, watermelon, and grapes? = Weekly

Or Other fruits such as fig, muskmelon, watermelon, and grapes? = Monthly

Q31 What other fruits do/did you eat?

▢ fig (4)
▢ grapes (5)
▢ muskmelon (6)
▢ watermelon (7)
▢ others (8)

Q32 Leaf vegetables such as lettuce, chicory, endive, turnip leaves, spinach, celery, purslane, cabbage, broccoli, brussel sprouts, cauliflower, beet leaves, fennel, arugula, green parts (leaves) of onions, and artichokes?

- Never (1)
- Daily (2)
- Weekly (3)
- Monthly (4)

Display This Question:

If Leaf vegetables such as lettuce, chicory, endive, turnip leaves, spinach, celery, purslane, cabba… = Daily

Or Leaf vegetables such as lettuce, chicory, endive, turnip leaves, spinach, celery, purslane, cabba… = Weekly

Or Leaf vegetables such as lettuce, chicory, endive, turnip leaves, spinach, celery, purslane, cabba… = Monthly

Q33 What leaf vegetables do/did you eat?

▢ arugula (4)
▢ beet leaves (5)
▢ broccoli (6)
▢ brussel sprouts (7)
▢ cabbage (8)
▢ cauliflower (9)
▢ celery (10)
▢ chicory (11)
▢ endive (12)
▢ fennel (13)
▢ green onions (14)
▢ lettuce (15)
▢ purslane (16)
▢ spinach (17)
▢ turnip leaves (18)
▢ artichokes (19)
▢ others (20)

Q34 Root vegetables such as carrot, turnip, garlic, beetroots, parsnip roots, parsley root, radish, onion, sacred lotus, taro, yam, sweet potato, potato?

- Never (1)
- Daily (2)
- Weekly (3)
- Monthly (4)

Display This Question:

If Root vegetables such as carrot, turnip, garlic, beetroots, parsnip roots, parsley root, radish, o… = Daily

Or Root vegetables such as carrot, turnip, garlic, beetroots, parsnip roots, parsley root, radish, o… = Weekly

Or Root vegetables such as carrot, turnip, garlic, beetroots, parsnip roots, parsley root, radish, o… = Monthly

Q35 What root vegetables do/did you eat?

▢ beetroots (5)
▢ carrot (6)
▢ garlic (7)
▢ onion (9)
▢ parsley roots (10)
▢ parsnip roots (11)
▢ potato (12)
▢ radish (13)
▢ sacred lotus (14)
▢ sweet potato (15)
▢ taro (16)
▢ turnip (18)
▢ yam (19)
▢ others (20)

Q88 Other vegetables such as cucumber, gourd, asparagus, leek, cherry tomato, garden tomatoes, green bell pepper, Italian sweet red pepper, orange bell pepper, yellow bell pepper, eggplant, okra?

- Never (1)
- Daily (2)
- Weekly (3)
- Monthly (4)

Display This Question:

If Other vegetables such as cucumber, gourd, asparagus, leek, cherry tomato, garden tomatoes, green… = Daily

Or Other vegetables such as cucumber, gourd, asparagus, leek, cherry tomato, garden tomatoes, green… = Weekly

Or Other vegetables such as cucumber, gourd, asparagus, leek, cherry tomato, garden tomatoes, green… = Monthly

Q89 What other vegetables do/did you eat?

▢ asparagus (4)
▢ cherry tomatoes (5)
▢ cucumber (6)
▢ gourd (7)
▢ eggplant (8)
▢ garden tomatoes (9)
▢ green bell pepper (10)
▢ Italian sweet red pepper (11)
▢ leek (12)
▢ okra (13)
▢ orange bell pepper (14)
▢ yellow bell pepper (15)
▢ others (16)

Q36 Beans and peas such as green beans, yellow wax bean, common bean, broad bean, Mung bean, black-eyed pea (cowpea), yardlong bean, scarlet bean, lima bean, common pea, chickpeas, carob beans, soybean?

- Never (1)
- Daily (2)
- Weekly (3)
- Monthly (4)

Display This Question:

If Beans and peas such as green beans, yellow wax bean, common bean, broad bean, Mung bean, black-ey… = Daily

Or Beans and peas such as green beans, yellow wax bean, common bean, broad bean, Mung bean, black-ey… = Weekly

Or Beans and peas such as green beans, yellow wax bean, common bean, broad bean, Mung bean, black-ey… = Monthly

Q37 What beans and peas do/did you eat?

▢ black-eyed pea (cowpea) (4)
▢ broad bean (5)
▢ carob beans (6)
▢ chickpeas (7)
▢ common beans (8)
▢ common peas (9)
▢ green beans (10)
▢ lima bean (11)
▢ Mung bean (12)
▢ scarlet beans (13)
▢ soybeans (14)
▢ yellow wax beans (15)
▢ yardlong bean (16)
▢ others (17)

Q38 Nuts and seeds such as almond, pecan nut, Colorado pinyon (pine nuts), Brazil nuts, Macadamia nuts, pistachio, hazel nuts, walnut?

- Never (1)
- Daily (2)
- Weekly (3)
- Monthly (4)

Display This Question:

If Nuts and seeds such as almond, pecan nut, Colorado pinyon (pine nuts), Brazil nuts, Macadamia nut… = Daily

Or Nuts and seeds such as almond, pecan nut, Colorado pinyon (pine nuts), Brazil nuts, Macadamia nut… = Weekly

Or Nuts and seeds such as almond, pecan nut, Colorado pinyon (pine nuts), Brazil nuts, Macadamia nut… = Monthly

Q39 What nuts and seeds do/did you eat

▢ almond (4)
▢ Brazil nuts (5)
▢ Colorado pinyon (pine nuts) (6)
▢ hazel nuts (7)
▢ Macadamia nuts (8)
▢ pecan nut (9)
▢ pistachio (10)
▢ walnuts (11)
▢ others (12)

Q40 Spices and herbs such as rosemary, thyme, coriander, ginger, sweet basil, dill, parsley, sage, rhubarb, saffron, chives, Chinese mustard, sweet marjoram, garden cress, sweet bay, turmeric, alfalfa, peppermint (including their extracted essential oils used as food additives and flavoring)?

- Never (1)
- Daily (2)
- Weekly (3)
- Monthly (4)

Display This Question:

If Spices and herbs such as rosemary, thyme, coriander, ginger, sweet basil, dill, parsley, sage, rh… = Daily

Or Spices and herbs such as rosemary, thyme, coriander, ginger, sweet basil, dill, parsley, sage, rh… = Weekly

Or Spices and herbs such as rosemary, thyme, coriander, ginger, sweet basil, dill, parsley, sage, rh… = Monthly

Q41 What spices and herbs do/did you eat?

▢ alfalfa (4)
▢ Chinese mustard (5)
▢ chives (6)
▢ coriander (7)
▢ dill (8)
▢ garden cress (9)
▢ ginger (29)
▢ parsley (10)
▢ peppermint (11)
▢ rhubarb (12)
▢ rosemary (13)
▢ saffron (14)
▢ sage (15)
▢ sweet basil (16)
▢ sweet bay (17)
▢ sweet marjoram (18)
▢ thyme (19)
▢ turmeric (20)
▢ others (21)

Q42 Cereals/grains such as or using buckwheat, barley?

- Never (1)
- Daily (2)
- Weekly (3)
- Monthly (4)

Display This Question:

If Cereals/grains such as or using buckwheat, barley? = Daily

Or Cereals/grains such as or using buckwheat, barley? = Weekly

Or Cereals/grains such as or using buckwheat, barley? = Monthly

Q43 What buckwheat and barley food do/did you eat? ()

Q44 Other foods such as mushroom?

- Never (1)
- Daily (2)
- Weekly (3)
- Monthly (4)

Display This Question:

If Other foods such as mushroom? = Daily Or Other foods such as mushroom? = Weekly

Or Other foods such as mushroom? = Monthly

Q45 What other foods do/did you eat?

▢ mushroom (4)
▢ Others (6)

Q46 Please tell us how frequently you ate/drank fermented foods during this pandemic to prevent catching COVID-19 or to facilitate recovery from COVID-19, such as sauerkraut, pickles, kefir, tempeh, natto, kombucha, miso, kimchi, yogurt? (check one answer that best describes the frequency of your use of the food category)

- Never (1)
- Daily (2)
- Weekly (3)
- Monthly (4) Display This Question:

If Please tell us how frequently you ate/drank fermented foods during this pandemic to prevent catch… = Daily

Or Please tell us how frequently you ate/drank fermented foods during this pandemic to prevent catch… = Weekly

Or Please tell us how frequently you ate/drank fermented foods during this pandemic to prevent catch… = Monthly

Q47 What fermented foods/drinks do you eat/drink?

▢ kefir (4)
▢ kimchi (5)
▢ kombucha (6)
▢ miso (7)
▢ natto (8)
▢ pickles (9)
▢ sauerkraut (10)
▢ tempeh (11)
▢ yogurt (12)
▢ others (13)

Display This Question:

If Please tell us how frequently you ate/drank fermented foods during this pandemic to prevent catch… = Daily

And Please tell us how frequently you ate/drank fermented foods during this pandemic to prevent catch… = Weekly

And Please tell us how frequently you ate/drank fermented foods during this pandemic to prevent catch… = Monthly

Q48 What food materials/ingredients are they made from?

Q49 Please tell us how frequently you took **<u>herbal medicines</u>** during this pandemic <u>**to prevent catching COVID-**</u> **<u>19 or facilitate recovery from COVID-19.</u>** (check one answer that best describes the frequency of your use of the food category)

- Never (1)
- Daily (2)
- Weekly (3)
- Monthly (4)

Display This Question:

If Please tell us how frequently you took herbal medicines during this pandemic to prevent catching… = Daily

Or Please tell us how frequently you took herbal medicines during this pandemic to prevent catching… = kly

Or Please tell us how frequently you took herbal medicines during this pandemic to prevent catching… = Monthly

Q50 Which herbal medicine(s) did you take?

Q51 Please tell us how frequently you took any **<u>supplements</u>** like vitamins, CBD pills, turmeric pills, propolis that you used during this pandemic **<u>to prevent catching COVID-19 or facilitate recovery from COVID-19.</u>** (check one answer that best describes the frequency of your use of the food category)

- Never (1)
- Daily (2)
- Weekly (3)
- Monthly (4)

Display This Question:

If Please tell us how frequently you took any supplements like vitamins, CBD pills, turmeric pills,… = Daily

Or Please tell us how frequently you took any supplements like vitamins, CBD pills, turmeric pills,… = Weekly

Or Please tell us how frequently you took any supplements like vitamins, CBD pills, turmeric pills,… = Monthly

Q52 Which supplement(s) did you take?

Q53 Please tell us how frequently you used some **<u>essential oils</u>** (include different delivery methods such as containers, diffusers, candles, massage, or other aromatherapeutic delivery methods) **<u>to prevent catching COVID-19.</u>** (check one answer that best describes the frequency of your use of the food category)

- Never (1)
- Daily (2)
- Weekly (3)
- Monthly (4)

Display This Question:

If Please tell us how frequently you used some essential oils (include different delivery methods su… = Daily

Or Please tell us how frequently you used some essential oils (include different delivery methods su… = Weekly

Or Please tell us how frequently you used some essential oils (include different delivery methods su… = Monthly

Q54 What type of smells/odorants?

Display This Question:

If Please tell us how frequently you used some essential oils (include different delivery methods su… = Daily

And Please tell us how frequently you used some essential oils (include different delivery methods su… = Weekly

And Please tell us how frequently you used some essential oils (include different delivery methods su… = Monthly

Q55 What delivery method(s) do you use?

▢ containers (4)
▢ diffuser (5)
▢ candles (6)
▢ massage (7)
▢ others (8)

Q56 Please tell how frequently you used some **<u>essential oils</u>** (include different delivery methods such as containers, diffusers, candles, massage, or other aromatherapeutic delivery methods) **<u>to facilitate recovery from COVID-19.</u>** (check one answer that best describes the frequency of your use of the food category)

- Never (1)
- Daily (2)
- Weekly (3)
- Monthly (4)
- Did not get sick by COVID-19 (5)

Display This Question:

If Please tell how frequently you used some essential oils (include different delivery methods such… = Daily

Or Please tell how frequently you used some essential oils (include different delivery methods such… = Weekly

Or Please tell how frequently you used some essential oils (include different delivery methods such… = Monthly

Q57 What type of smells/odorants?

Display This Question:

If Please tell how frequently you used some essential oils (include different delivery methods such… = Daily

Or Please tell how frequently you used some essential oils (include different delivery methods such… = Weekly

Or Please tell how frequently you used some essential oils (include different delivery methods such… = Monthly

Q58 What delivery method(s) do you use?

▢ container (4)
▢ diffuser (5)
▢ candle (6)
▢ massage (7)
▢ others (8)

Q59 Please tell us if there are **anything else** (for example, air freshener, perfume) that you use daily during this pandemic **to prevent catching COVID-19 or facilitate recovery from COVID-19.**

Display This Question:

If If Please tell us if there are anything else (for example, air freshener, perfume) that you use dail… Text Response Is Not Empty

Q60 How often did you use it/them?

- Never (1)
- Daily (2)
- Weekly (3)
- Monthly (4)

Q61 Overall, are there some prescribed medicines, generic medicines, herbal medicines, home remedies, supplements, food ingredients, fluids, essential oils, or others that you think are effective in treating sickness and recovering from COVID-19 symptoms?

- Yes (1)
- No (2)
- Did not get COVID-19 (3)
- Did not try anything (4)

Display This Question:

If Overall, are there some prescribed medicines, generic medicines, herbal medicines, home remedies,… = Yes

Q62 If yes, please describe up to the top three:

- 1. What was it? (1)
- 1. For what symptom? (2)
- 2. What was it? (3)
- 2. For what symptom? (4)
- 3. What was it? (5)
- 3. For what symptom? (6)

Display This Question:

If Overall, are there some prescribed medicines, generic medicines, herbal medicines, home remedies,… = No

Q63 If No, is there anything that you heard that helps and you tried, but did it not work? Please describe up to three:

- 1. What was it? (1)
- 1. For what symptom? (2)
- 2. What was it? (3)
- 2. For what symptom? (4)
- 3. What was it? (5)
- 3. For what symptom? (6)

Q64 What year were you born?

Q65 What is your gender?

- Male (1)
- Female (2)
- Non-binary / third gender (3)
- Prefer not to say (4) Q66 What is your ethnicity?

Q67 Your current location.

- City (1)
- Country (2)

Q68 Do you consider the answers you provided here to be strongly influenced by a specific (or several specific) country/culture(s)?

- Yes (1)
- No (2)

Display This Question:

If Do you consider the answers you provided here to be strongly influenced by a specific (or several… = Yes

Q69 If yes, which country or culture?

Q70 Do you smoke?

- Yes (1)
- No (2)

Q71 Do you vape?

- Yes (1)
- No (2)

Display This Question:

If Do you vape? = Yes

Q72 If yes, which flavor(s) do you add?

Q73 Did you have pre-existing health conditions pre-pandemic?

- Yes (1)
- No (2)

Display This Question:

If Did you have pre-existing health conditions pre-pandemic? = Yes

Q74 Which health conditions?

Q75 Is there anything else that you would like to share about how best to treat Covid-19 symptoms?

- Yes (4)
- No (5)

Q76 Lastly, did your eating/drinking habits change from pre-pandemic? If yes, explain. Did you stop eating certain items? Did you start eating certain items more?

- Yes, started to eat/drink more XX: Please tell me what you started to eat/drink more (4)
- Yes, started not to eat/drink YY: Please tell us what you started not to eat/drink or eat/drink less (5)
- No (6)

End of Block: Block 2

